# Effectiveness of Lesser Known Herbal Sedatives for Insomnia: A Systematic Review and Meta-Analysis

**DOI:** 10.64898/2026.03.23.26349099

**Authors:** Muhammad Awais Paracha, Sania Abdul Jalil Khan, Rida Zarkaish, Faryal Fazal, Muhammad Danish Khan, Mahnoor Ahmad

**Author notes:** **Corresponding Author:** Muhammad Awais Paracha. Clinical Trial Number: Not applicable. Systematic Review Registration: PROSPERO CRD420251101795.

## Abstract

**Background:** Insomnia is a major public health problem affecting an estimated 852 million adults worldwide. Current pharmacological treatments, including benzodiazepines and Z-drugs, carry serious risks of dependency, cognitive impairment, and adverse events. These limitations have driven growing interest in complementary and alternative therapies, particularly herbal sedatives, which are perceived as natural and safer. However, evidence on their safety and efficacy remains insufficient and patchy.

**Objective:** This review evaluated the effectiveness of lesser known herbal sedatives for insomnia.

**Methods:** The protocol was registered with PROSPERO (CRD420251101795). Eligibility was defined using the PICO framework: **Population:** adults aged ≥18 years with insomnia; **Interventions:** Passiflora incarnata, Hawthorn, Melissa officinalis, Chamomile, Viola odorata, Nelumbo nucifera, Rhodiola rosea, and Eschscholtzia californica. **Comparators:** placebo or usual care; **Primary and Secondary Outcomes:** sleep quality (Pittsburgh Sleep Quality Index, Insomnia Severity Index, Epworth Sleepiness Scale), sleep duration, and sleep latency. Databases and registers were searched from January 2005 to July 2025. Randomized controlled trials, nonrandomized controlled trials, clinical trials, and observational studies were included. Five reviewers independently screened studies. Data extraction used a structured Excel spreadsheet. Risk of bias was assessed using RoB 2.0 for randomized trials and ROBINS-I V2 for nonrandomized studies. Random-effects meta-analyses (DerSimonian and Laird) were conducted in RevMan. Narrative synthesis followed SWiM guidelines.

**Results:** From 1,294 records, 32 studies met eligibility criteria. Meta-analysis of 23 RCTs demonstrated a statistically significant pooled effect favouring herbal sedatives (SMD −0.77, 95% CI −1.14 to - 0.40, p=0.0001), with substantial heterogeneity (I²=92%). Subgroup analysis showed larger effects for chamomile (SMD −1.06) and Melissa officinalis (SMD −0.66). Most RCTs had high overall risk of bias; nonrandomized studies predominantly had critical risk of bias.

**Conclusions:** This systematic review provides preliminary evidence that several herbal sedatives, particularly chamomile and Melissa officinalis, may improve insomnia-related outcomes. However, methodological weaknesses, high risk of bias, and substantial heterogeneity limit evidence strength. Future research requires standardized extracts, large multicentre RCTs, and extended follow-up.

**Article Summary:** *Strengths and limitations of this study:*

- This systematic review followed a pre-registered protocol (PROSPERO CRD420251101795) and adhered to PRISMA 2020 guidance, with comprehensive searches of multiple databases and trial registers without language restrictions.
- Six reviewers independently performed study selection, data extraction, and risk of bias assessment using Cochrane RoB 2.0 for randomised trials and ROBINS-I for non-randomised studies, with disagreements resolved through discussion or arbitration.
- The review included both meta-analysis of randomised controlled trials and narrative synthesis of non-randomised studies following Synthesis Without Meta-analysis (SWiM) guidelines, providing a comprehensive evaluation of the available evidence.
- Despite extensive searches, identifying single herbal interventions for several botanicals proved challenging, with some herbs appearing only in non-randomised trials or combination products, preventing definitive conclusions about their individual efficacy.
- The evidence is limited by substantial statistical heterogeneity (I² = 92%), high or critical risk of bias in most included studies, and statistically significant publication bias detected by Egger’s test (p = 0.0217), which limits confidence in the pooled effect estimates. Using the GRADE framework, the overall certainty of evidence was rated as moderate

## Background

Insomnia is widely recognized as a major public health problem that is associated with serious health consequences, such as cardiovascular morbidity, mood disorders, cognitive impairment, and lower quality of life, if left untreated for a long period of time [1, 2] According to recent estimates, approximately 16.2% of the adult population, i.e., 852.3 million people, experience insomnia worldwide, and nearly 415 million have severe symptoms [1]. Prevalence rates are higher in females of all age groups [1]. In recent years, modern lifestyle habits, specifically the excessive use of digital media, have become increasingly associated with sleep disorders, increasing the relevance of insomnia and current issues for investigation. Growing evidence suggests that the excessive use of smartphones, tablets, and other digital devices, particularly during evening hours, is linked to delayed bedtime, a lack of total sleep, poor sleep quality, and daytime sleepiness [3, 4, 5]. Exposure to bright screen light in the evening may postpone endogenous melatonin release and disrupt circadian rhythms, which leads to sleep latency [6.7]. Electronic devices further suppress the production of melatonin, which is an important hormone that facilitates sleep onset and maintenance due to the emission of light of short wavelengths [8, 9]. In addition, physiological arousal due to the use of social media or engaging in interactive online entertainment at night may lead to problems falling or staying asleep [10]. These interrelated physiological and behavioural mechanisms have substantially contributed to contemporary trends in insomnia research.

The current approaches for treating insomnia involve both pharmacological and nonpharmacological methods. The American Academy of Sleep Medicine (AASM) suggested that Cognitive Behavioral Therapy for Insomnia (CBT-I) should be used as the first-line treatment because of its effectiveness in increasing sleep initiation, maintenance and long-term sleep outcomes [11]. However, the use of CBT-I is limited by its low availability, high cost and lack of trained therapists. Benzodiazepines and nonbenzodiazepine hypnotics (Z-drugs) are widely prescribed pharmacological treatments for insomnia; have serious side effects, such as dependency, addiction drug tolerance, cognitive impairment, daytime sedation and poor long-term health outcomes; and, in severe cases, fatal events (e.g., carbon monoxide poisoning, drowning, falls, hypothermia, motor vehicle collisions, and apparent suicide), as well as nonfatal injuries, such as accidental overdoses, gunshot wounds, burns, and self-inflicted harm [12,13]. Z-drugs also have boxed warnings issued by the U.S. Food and Drug Administration (FDA) because they have potentially harmful sleep-related behaviours such as sleep walking and sleep driving [14]. Melatonin, melatonin receptor agonists, and sedating antidepressants (e.g., Trazodone) are also used in clinical practice, even though their effectiveness as agents in treating chronic insomnia is inconclusive [15, 16]. These limitations of contemporary pharmacological and nonpharmacological treatments have led to a growing interest in complementary and alternative therapies (CAMs) for insomnia [17].

Among the widely used CAM therapies are herbal sedatives, which are perceived as natural and safer, leading many individuals to use them as independent agents or alongside other conventional therapies; however, the evidence on their safety is insufficient [18]. The use of herbal supplements in Ayurveda and traditional Chinese medicine has a long history and remains popular even today [19]. Similarly, the worldwide market of herbal sedatives has grown significantly over the past few years [20]. Despite their widespread use, herbal preparations do not have a standardized dosage, and their regulation, particularly in some parts of the world, such as the United States and Europe, is less stringent than that of pharmaceutical drugs [21, 22]. Although research interest in herbal sedatives has recently increased, interest in this topic remains patchy in the scientific literature. A systematic review and meta-analysis of randomized controlled trials revealed moderate effects of valerian on sleep latency and sleep quality [23]. Similarly, chamomile has shown potential benefits in improving sleep quality [24].

In contrast, other commonly used herbal sedatives, such as Passion flower (Passiflora incarnata), Lemon balm (Melissa officinalis), Hawthorn (Crataegus oxyacantha), Sweet violet (Viola odorata), Lotus (*Nelumbo nucifera*), California poppy (Eschscholtzia californica) and Rhodiola (*Rhodiola rosea*), have received little coverage in the literature [25, 26, 27, 28, 29, 30, 31, 32]. The existing studies on these lesser known herbal sedatives are limited, methodologically inconsistent and have small sample size [33, 34, 35]. Moreover, some herbs have dose- and formulation-dependent effects; for example, ***R.*** *rosea* can be stimulating rather than having sedative effects. Likewise, safety data for many other herbs, particularly alkaloid-containing herbs such as Eschscholtzia californica*, are* limited, with significant gaps in our knowledge of long-term effects, combinatory risks, and use in vulnerable populations such as pregnant or breastfeeding individual [36,37]. These gaps in the literature emphasize the need for a systematic evaluation of these lesser-known herbal sedatives used for insomnia and highlight the necessity of a rigorous systematic review to identify the efficacy and safety of these herbs used to treat sleep disorders. Therefore, this study aims to evaluate the effectiveness of these lesser known herbal sedatives for the treatment of insomnia and sleep disorders.

## Methods

### Registration and Protocol

The protocol was registered with the PROSPERO (International Prospective Register of Systematic Reviews) (registration number CRD420251101795). Available from: https://www.crd.york.ac.uk/PROSPERO/view/CRD420251101795.

### Eligibility criteria

The eligibility criteria were based on the Population, Intervention, Comparator, Outcome (PICO) framework, as suggested in the PRISMA 2020 expanded checklist [38]. The inclusion and exclusion criteria were developed to include relevant and high-quality studies.

### Population

Primary research studies on adults aged 18 years or above who experienced insomnia or sleep disorders were included. Studies with populations younger than 18 years, i.e., adolescents and children, were excluded to ensure consistency.

### Intervention

Eligible interventions consisted of herbal interventions of a prespecified group: passionflower (Passiflora incarnata), hawthorn (Crataegus spp.), lemon balm (Melissa officinalis), chamomile (Matricaria chamomilla), sweet violet (Viola odorata), lotus (*Nelumbo nucifera*), Rhodiola (*Rhodiola rosea*), and California poppy (Eschscholtzia californica) or a combination of these interventions. Interventions using homeopathic or allopathic agents or herbal agents not specified above were excluded.

### Comparator

Studies with the following comparators were eligible: placebo, usual care, cognitive behavioural therapy (CBT), or pharmacotherapy. This ensured that the estimated effects of the herbal interventions could be meaningfully interpreted relative to both active and inactive controls.

### Outcomes

The primary outcomes were sleep quality measured through validated tools such as the Pittsburgh Sleep Quality Index (PSQI), the Insomnia Severity Index (ISI) or the Epworth Sleepiness Scale (ESS), sleep duration in minutes or hours, and sleep latency, which were assessed from baseline to the final follow-up. The secondary outcomes included the frequency of night-time awakenings and quality of life.

### Study Design

Randomized controlled trials (RCTs), nonrandomized controlled trials, clinical trials, observational studies, and quasi-experimental studies were included if full-text articles with institutional access were available. Studies published only as conference abstracts or nonindexed articles without accessible full texts were excluded. Studies conducted in both clinical and community-based settings were eligible, and inclusion was not restricted by language, provided that an English translation was available utilizing language translation tools.

### Information Sources

A comprehensive search strategy was implemented to identify relevant studies. The following databases and sources were systematically searched until July 2025: the Cochrane Library via the official Cochrane platform, PubMed via the NCBI interface, EuroPMC, EBSCO host databases, Google Scholar, and Trial Registers (including ClinicalTrials.gov and the WHO ICTRP).

No restrictions were placed on publication language or status, although only studies published in the last twenty years, i.e., January 2005 to July 2025, were considered. Unpublished studies and gray literature were also searched in Open Thesis and Dissertations (OTAD). Additional sources included manual reference list searches of relevant included studies.

### Search strategy

The review team developed the search strategy using relevant keywords and was peer-reviewed by an experienced systematic reviewer to ensure its accuracy and completeness. The strategy was customized for each database and trial registries with their relevant search terms. To ensure transparency and reproducibility, the full line-by-line search string for each database is available in Appendix I. The search strategy

The following keywords were searched:

Insomnia, “sleep initiation and maintenance disorders,” “sleep wake disorders,” “sleep quality,” “sleep latency,” “passiflora incarnate or passionflower,” “crataegus or hawthorn,” “melissa officinalis or lemon balm,” “matricaria chamomilla or chamomile,” “viola odorata or sweet violet,” “nelumbo nucifera or lotus,” “rhodiola rosea or rhodiola,” “eschscholtzia californica or california poppy.”

### Selection process

The selection process adhered to the PRISMA 2020 guidance, with a two-stage screening approach:

1. **Title and Abstract Screening**: Five reviewers (MAP, RZ, SAJK, MDK and MA) independently screened the titles and abstracts of all retrieved records against the prespecified inclusion and exclusion criteria in a double-blinded manner. Studies that clearly did not meet the criteria were excluded at this stage for various reasons. Any disagreements among the reviewers were discussed until a consensus was reached. The sixth reviewer (FF) was consulted if consensus was not achieved, and the majority’s decision was considered final.
2. **Full-Text Screening**: Full-text articles of potentially eligible studies were retrieved and independently assessed in a double-blinded manner by three reviewers (MAP, RZ and SAJ). Each article was evaluated against the eligibility criteria. For all studies excluded at this stage, the reasons for exclusion were also recorded for transparency and eventual inclusion in the Reporting Items for Systematic Reviews and Meta-Analyses (PRISMA) flow diagram [38].
3. Rayyan AI was used without enabling AI-assisted ranking of articles during the screening process [39]. The authors of the studies were contacted for clarification or for additional information if needed. Articles requiring translation were translated with the assistance of tools such as Google Translate (https://translate.google.com) and EZ AI Translate (https://ezaitranslate.com/#document-translate). Translation was performed twice to ensure accuracy. The authors of the review have some basic proficiency in the Persian language, especially numerals, and the translated data were duly verified for articles translated from persion.

### Data collection process

Data extraction was conducted via a structured Microsoft Excel spreadsheet developed and pilot tested by the review team, adapted from the Cochrane data collection form to capture all relevant study characteristics and variables [40]. This approach ensured consistency and comprehensiveness across the included studies. Six reviewers (MAP, RZ, SAJ, FF, MDK and MA) independently extracted data from the full-text articles meeting the eligibility criteria.

### Data Items

**The following data items were extracted:**

- **Study characteristics**: Author(s), year, country, study design, setting, sample size.
- **Participant characteristics**: Inclusion/exclusion criteria, age, sex, and baseline sleep characteristics.
- **Intervention details**: Herbal agent, dosage, route of administration, duration, and frequency.
- **Comparator details**: Type of control (placebo, usual care, CBT, pharmacotherapy).
- **Primary outcomes**: Sleep quality (PSQI global score), insomnia severity (ISI), sleep quality, sleep duration, and sleep latency at all available follow-up time points.
- **Secondary outcomes**: Frequency of night-time awakening and quality of life.

For each outcome reported in the study, data were collected. If multiple time points were reported, data were extracted from baseline until the last follow-up. Assumptions regarding missing or unclear data were made conservatively.

### Risk of Bias Assessment

The methodological quality of the included studies was assessed via the Cochrane risk of bias tools appropriate for study design. For randomized controlled trials, we utilized the revised Cochrane Risk of Bias tool 2.0 for randomized trials (RoB 2.0) [41]. Four reviewers (MAP, RZ, SAJ and FF) independently performed risk of bias assessments twice, with disagreements resolved through discussion or arbitration. We recorded all judgments and supporting statements verbatim from the included studies via the official Microsoft Excel-based RoB 2.0 tool available from the Cochrane risk of bias website (https://www.riskofbias.info/welcome/rob-2-0-tool/current-version-of-rob-2. Overall risk of bias judgments were made according to the RoB 2 decision algorithm, categorizing studies as low, some concerns, or high risk of bias.

We also performed a thorough risk of bias assessment for nonrandomized control trials utilizing the ROBINS-I V2 instrument [42]. Two independent assessors (MAP, MDK) employed the ROBINS-I V2 tool and recorded direct quotations from the studies to rationalize each evaluation. The risk of bias levels for individual domains, i.e., low, moderate, serious, and critical, were assigned according to the ROBINS-I signalling questions. Any disagreements between reviewers were resolved through discussion or by involving a third arbitrator (MA). All outputs were captured via a macroenabled Microsoft Excel tool developed by Huang (2025) [43]. For a clear summary, the distribution of bias was visualized with robvis, creating standard traffic-light plots and summary plots for each domain [44].

### Effect Measures

For continuous outcomes (e.g., sleep quality, sleep duration, and sleep latency), the standardized mean difference (SMD) was used, depending on the consistency of measurement scales across studies. The SMD was preferred for outcomes measured with different validated scales (e.g., the PSQI vs. the ISI). Thresholds for interpreting the magnitude of effect were based on established guidelines (e.g., SMDs of 0.2, 0.5, and 0.8 interpreted as small, moderate, and large effects, respectively [45]).

### Synthesis methods

#### Eligibility for Synthesis

Studies were grouped for synthesis by study type (i.e., randomized vs. nonrandomized trials), intervention type, and outcome measures. Randomized controlled trials (RCTs) sufficiently similar in terms of PICO components were included in the meta-analyses. For nonrandomized controlled studies, narrative synthesis was performed.

#### Tabulation and Graphical Methods

The data are summarized in structured tables that display the study and participant characteristics, interventions, comparators, outcomes, and risk of bias. Forest plots were used to visually display the meta-analytic results, funnel plots were used to depict publication bias, and risk of bias summary figures were generated via the robvis app [44].

#### Statistical Synthesis

Random-effects meta-analyses were conducted via the DerSimonian and Laird method implemented in RevMan software. Confidence intervals for summary effects were calculated via Wald-type intervals. Narrative synthesis was conducted for nonrandomized controlled trials via the Synthesis without Meta-analysis (SWiM) guidelines [46].

### Grouping studies for synthesis

Studies were grouped to directly address the review’s objective to synthesize evidence for prespecified herbal interventions: Passionflower, Hawthorn, Lemon Balm, Chamomile, Sweet Violet, Lotus, Rhodiola, and California Poppy.

- Group 1: Single herbs: Studies evaluating a single botanical from the protocol list.
- Group 2: Multiple Herb: Combinations with Confounding Interventions. Studies where a prespecified herb was combined with an intervention not on the protocol list (e.g., valerian or melatonin). These studies are acknowledged but not synthesized to avoid confounding.

#### Standardized Metric and Synthesis Method

The primary synthesis method was vote counting on the basis of the direction of effect. For all outcomes except Lemoine et al., 2019 “Sleep Quality” (0–10 scale), a positive change, i.e., (pre minus post scores), indicates improvement. For Lemoine et al., 2019, a negative change indicates improvement. The effects were categorized as follows:

- Beneficial (B): Statistically significant improvement (p < 0.05) and/or a change meeting the minimal clinically important difference (MCID; e.g., a 6–8 point reduction on the ISI).
- Likely Beneficial (LB): A clear improvement in means that does not meet the full criteria for “Beneficial.”
- No change/unclear (NC/U): Insufficient data, minimal change, or findings of ambiguous clinical relevance

### Heterogeneity

Heterogeneity among the included RCTs was assessed via the I² statistic, with I² > 50% indicating substantial heterogeneity.

### Subgroup analysis

To explore potential sources of the anticipated heterogeneity and to investigate whether particular herbal preparations or modes of delivery might exert differential effects, we conducted prespecified subgroup analyses. Subgrouping was performed according to (1) type of herbal sedative (chamomile, Melissa officinalis, Passiflora, combination remedies, and sweet violet) and (2) mode of administration (oral versus intranasal). These subgroups were selected a priori on the basis of clinical relevance and the anticipated variation in pharmacological profiles across different botanicals.

For subgroup analyses by type of herbal sedative, studies were grouped according to the primary active botanical intervention. Studies evaluating chamomile in any form (capsule, tea, essential oil aromatherapy, or inhalation) were aggregated into a single chamomile subgroup. Similarly, studies reporting Melissa officinalis as the sole botanical intervention were grouped together, irrespective of formulation or dosage. Combination remedies were defined as any intervention containing two or more herbal constituents, regardless of whether they included prespecified botanicals. The sweet violet subgroup comprised studies evaluating Viola odorata as a standalone intervention. For subgroup analysis by mode of administration, studies were dichotomized into oral administration (including capsules, tablets, teas, and liquid extracts taken by mouth) and intranasal administration (including essential oil inhalation and aromatherapy). Statistical heterogeneity within each subgroup was assessed using the I² statistic. Between-subgroup differences were evaluated using a chi-squared test, with a significance threshold of p < 0.10, as recommended for subgroup analyses in systematic reviews. All subgroup analyses were conducted within the random-effects model framework using RevMan, with the DerSimonian and Laird method applied to each subgroup independently.

### Sensitivity analysis

We conducted a series of sensitivity analyses to assess the robustness and stability of the pooled effect estimates and to investigate the influence of methodological decisions on the overall findings. These analyses were prespecified to evaluate whether the observed treatment effect was driven by specific study characteristics or by the inclusion of studies at a high risk of bias.

First, to assess the influence of combination remedies on the overall effect estimate, we performed a sensitivity analysis restricting the meta-analysis to studies evaluating single-herb interventions only. Studies that administered two or more herbal constituents concurrently were excluded from this analysis, and the pooled effect size was recalculated using the same random-effects model. This approach allowed us to determine whether the inclusion of multi-herb formulations, which may involve complex pharmacokinetic interactions, diluted or amplified the overall treatment effect.

Second, to investigate the impact of the mode of administration on the pooled effect size and the degree of between-study heterogeneity, we conducted a sensitivity analysis excluding studies that employed intranasal administration (aromatherapy or inhalation). The rationale for this exclusion was the substantial difference in bioavailability and pharmacological exposure between oral and intranasal routes, which could contribute to the high heterogeneity observed in the primary analysis. The pooled effect size and I² statistic were recalculated after the exclusion of intranasal studies, and the results were compared with those of the primary analysis.

All sensitivity analyses were conducted using the same random-effects model (DerSimonian and Laird method) implemented in RevMan, and the results were compared with the primary analysis with respect to both the magnitude and the statistical significance of the pooled effect, as well as the degree of statistical heterogeneity (I²). The findings of these analyses were interpreted to inform the overall robustness of the review’s conclusions.

### Reporting Bias

The risk of reporting bias for publication was calculated via R funnel plots. Egger’s test was performed for the formal assessment of small study effects. Selective outcome reporting within studies was evaluated as part of the risk of bias assessment.

### Certainty assessment

The overall certainty of the body of evidence was judged by two reviewers (MAP and MDK) working separately. To perform this evaluation, we used the GRADE (Grading of Recommendations, Assessment, Development, and Evaluations) system as implemented in the GRADEpro Guideline Development Tool, which adheres to the methodological standards set by Cochrane [47, 48, 49]. The step-by-step assessment was performed for all five core domains, i.e., risk of bias in the included studies, the consistency of findings across studies, the precision of the effect estimates, the directness of the evidence to the research question, and the likelihood of publication bias that can impact confidence in the results for each of our primary outcomes. On the basis of this assessment, each major outcome was assigned a certainty rating of high, moderate, low, or very low. Ratings were adjusted upwards or downwards where justified, with changes documented in the evidence summary table footnotes.

## Results

### Study Selection

A systematic search of databases and registries between June and July 2025 yielded 1,294 records. After removing 75 duplicates, we screened the titles and abstracts of the remaining 1,219 records, which led to the exclusion of 1,177 articles. The full texts of the remaining 42 studies were assessed for eligibility, resulting in the exclusion of 10 articles because of the following: wrong population (n=4), wrong outcome (n=1), wrong intervention (n=1), wrong study design (n=1), and wrong publication type (n=3). Consequently, 32 studies satisfied all eligibility criteria and were included in the final synthesis. The study selection process, which followed the Preferred Reporting Items for Systematic Reviews and Meta-Analyses (PRISMA) methodology [38], is illustrated in Figure 1.

**Figure 1:**
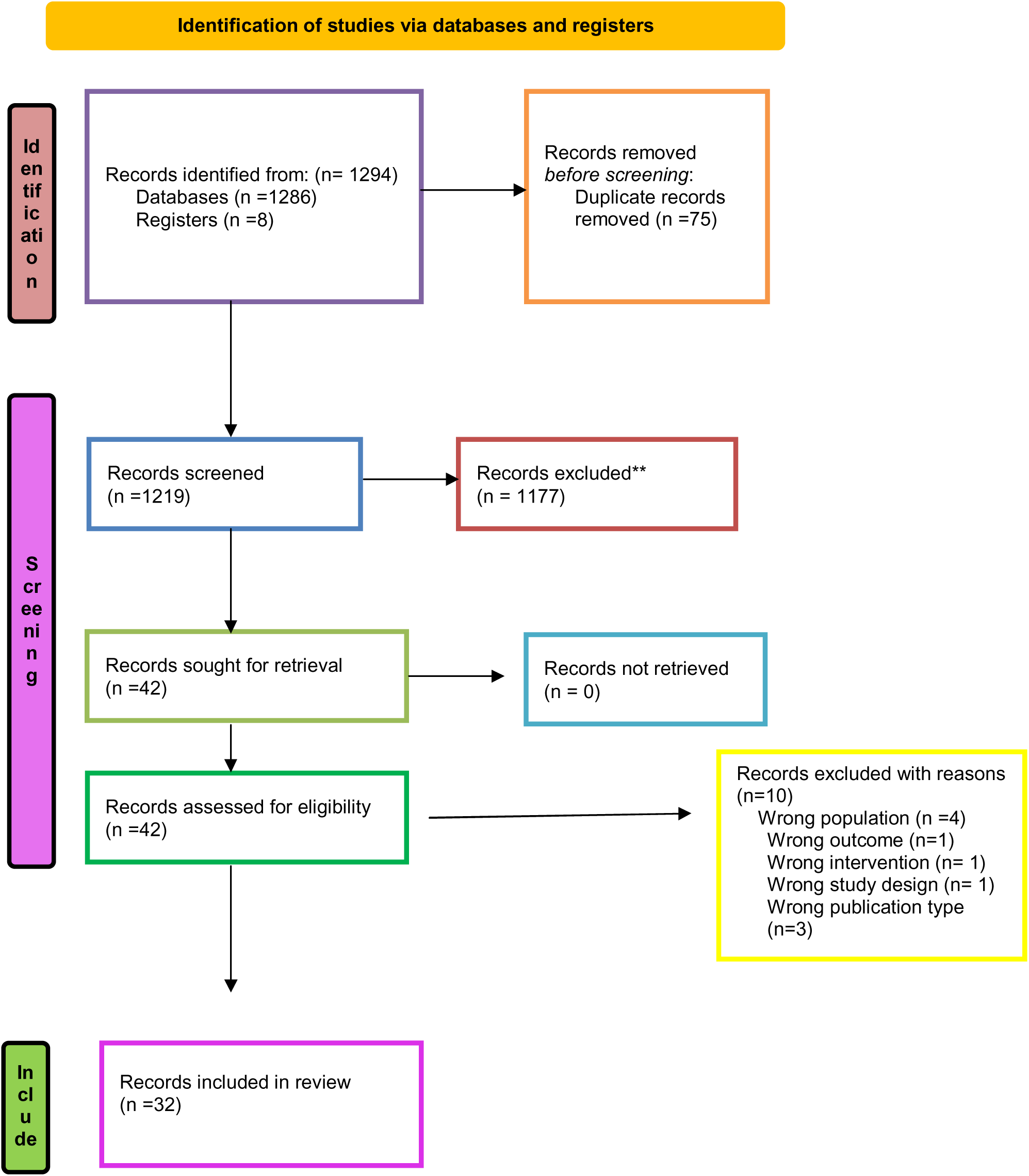
Preferred Reporting Items for Systematic Reviews and Meta-Analyses (PRISMA) flow diagram.

### Study characteristics

The general characteristics of the 23 RCTs are presented in Table 1: Appendix II Study characteristics of the RCTs. These studies were conducted in India, Iran, Indonesia, the United States of America, Thailand, Peru, Taiwan, Korea, Australia, Spain, Italy, Colombia and Germany between 2010 and 2025 and included 1541 participants. The participants’ ages ranged from 18--80 years, with sample sizes varying from 30--132 participants. The sleep-related outcomes were assessed via validated tools such as the Pittsburgh Sleep Quality Index (PSQI), the Insomnia Severity Index (ISI), sleep diaries, and additional stress, anxiety, or quality-of-life measures. Nineteen studies (82.6%) used the PSQI, followed by the ISI, which was used in 4 studies (17.4%). Subjective sleep quality (SQ) and the modified Athens Insomnia Scale (mAIS) were each used in one study, accounting for 4.3% of the RCTs. Depending on the intervention, the follow-up duration varied from 1 week to 12 weeks. The chamomile (Matricaria chamomilla L.) was evaluated in 10 RCTs in different modes of administration. These include chamomile extract capsules, chamomile high-grade extracts, chamomile essential oil aromatherapy, chamomile tea, chamomile oil inhalation, and combined interventions such as bergamot plus chamomile tea. The intervention duration varied from 2 to 8 weeks. Four RCTs utilized Passiflora incarnate, passion flower in oral capsules and herbal tea, with treatment durations ranging from 1--6 weeks. A total of 4 RCTs used Melissa officinalis, commonly known as lemon balm, in different modes, sometimes alone or in combination. The treatment durations ranged from 2 to 4 weeks. Five RCTs utilized combinations of Melissa officinalis, sweet violet, and lemon balm along with other multiple botanical agents. The treatment durations ranged from 2--6 weeks, and different trials were performed.

**Table 1: Appendix II: Study characteristics of the RCTs**

The study characteristics of the 9 non-randomized controlled trials as presented in Table 2: Appendix II: Study characteristics of nonrandomized studies. These studies were conducted in France, India, Iran, Italy, the Republic of Korea, Poland, and Thailand and included 369 participants in total. The sample size varied from 20 to 100 individuals aged between 16 and 90 years. Across trials, treatment durations ranged from 2--8 weeks, with outcomes commonly assessed via tools such as the Insomnia Severity Index (ISI), Pittsburgh Sleep Quality Index (PSQI), sleep diaries, and measures of depressive and anxiety symptoms, as well as quality of life. Five out of the 9 non-RCTS focused on single-herb interventions (e.g., chamomile, lemon balm, sweet violet, Melissa officinalis), whereas 4 out of the 9 evaluated multiherbal combinations that included botanical/non-botanical blends.

**Table 2: Appendix II: Study characteristics of nonrandomized studies Risk of bias assessment**

We used the Cochrane risk of bias tool (RoB 2.0) to determine the risk of bias of the 21 parallel-arm randomized controlled trials. Most studies reported a high overall risk of bias. This group comprises Abdullahzadeh et al. (2017) [50], Abdollahzadeh and Naji (2014) [52], Abbasinia et al. (2016) [53], Alvarado-Garcia et al. (2024) [55], Chang and Chen (2016) [56], Adib Hajbaghery et al. (2017) [57], Deepa et al. (2024) [58], Renosih et al. (2025) [59], Harit et al. (2024) [60], and Lee et al. (2019) [61]. In these trials, Domain 2 (deviations of the intended interventions) and Domain 4 (measures the outcome) were the main sources of bias. A high rating in Domain 2 usually means that the participants and personnel were not blinded and that the analysis did not consider the possibility of this bias. A high rating of Domain 4 indicated that the outcome was not blinded and that assessment of the outcome could be influenced by familiarity with the intervention. Domain 3 (missing outcome data) was also an issue that was frequently expressed, and it also posed some concerns with respect to dropouts and the completeness of the data.

The second group of studies was rated as having ‘some concerns’ regarding the overall risk of bias. Examples of this category are Jarukitsakul et al. (2025) [54]; Ranjbar et al. (2018a) [67]; Taherzadeh et al. (2021) [69]; and Gutierrez-Romero et al. (2024) [71]. Problems were common in Domain 1 (randomization process), in which the method or allocation concealment might not have been sufficient, and in Domain 5 (selection of the reported result), in which it was possible to selectively report the outcomes.

Zick et al. (2011) [51] was the only study that was rated as having a low risk of bias in all domains, which made their overall assessment low. The trial involved sound procedures, such as adequate randomization, a low risk of bias due to missing data, blind estimation of outcomes, and a preplanned, nonselective analysis strategy. Figure 2 shows risk of bias assessment parallel arm trials

**Figure 2:**
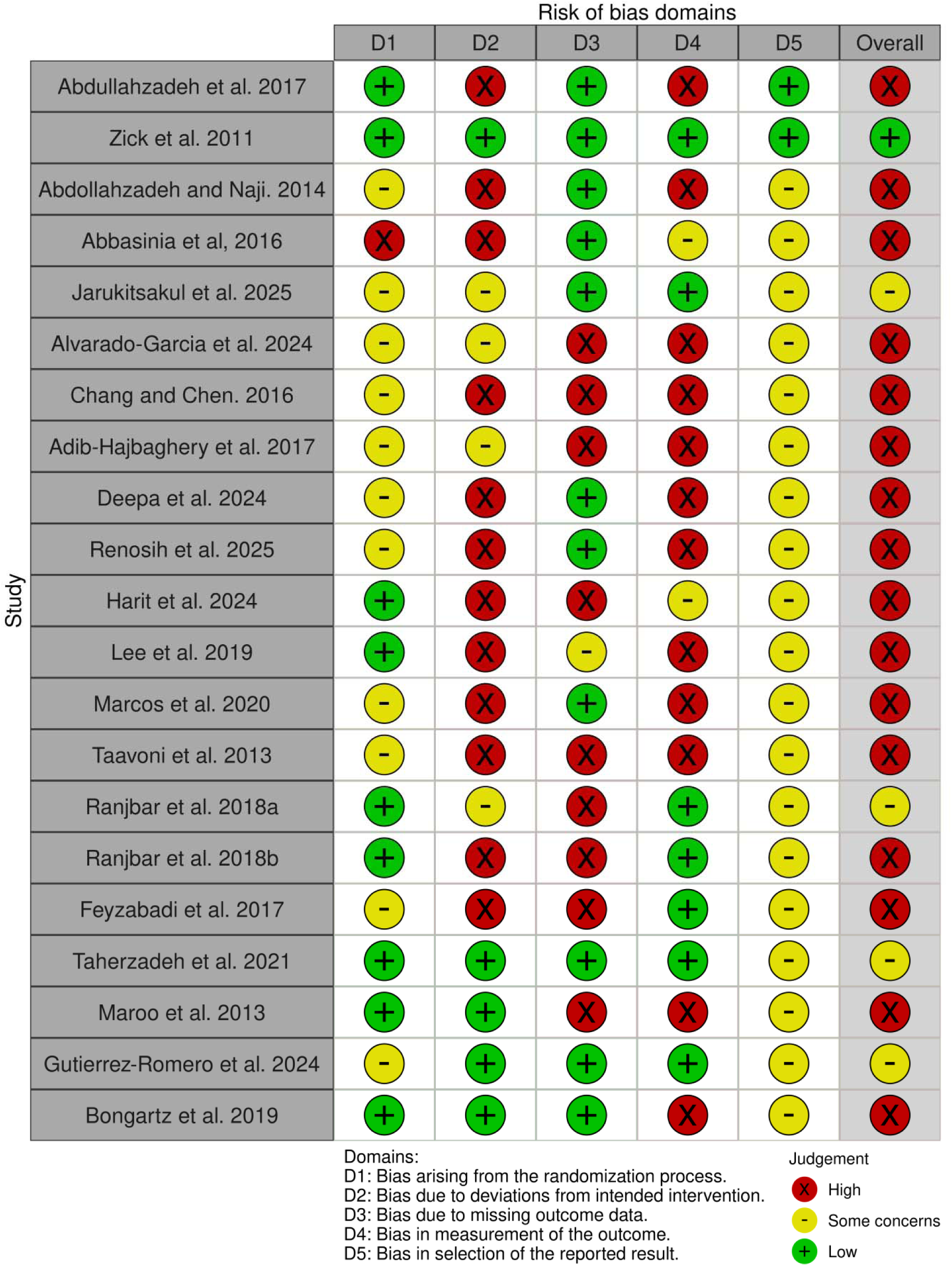
Risk of bias assessment parallel arm trials.

### Risk of Bias Assessment Crossover Trials

The risks of bias assessment of two crossover trials, i.e., Ngan and Conduit (2011) [62] and Pierre et al. (2024) [65], were rated as having a high overall risk of bias, mainly because Domain 4 (measurement of the outcome) had a high risk associated with the absence of blinding in the outcome assessment. In both, certain issues were observed in Domain 5 (selection of reported results). Figure 3 shows risk of bias assessment crossover trial

**Figure 3:**
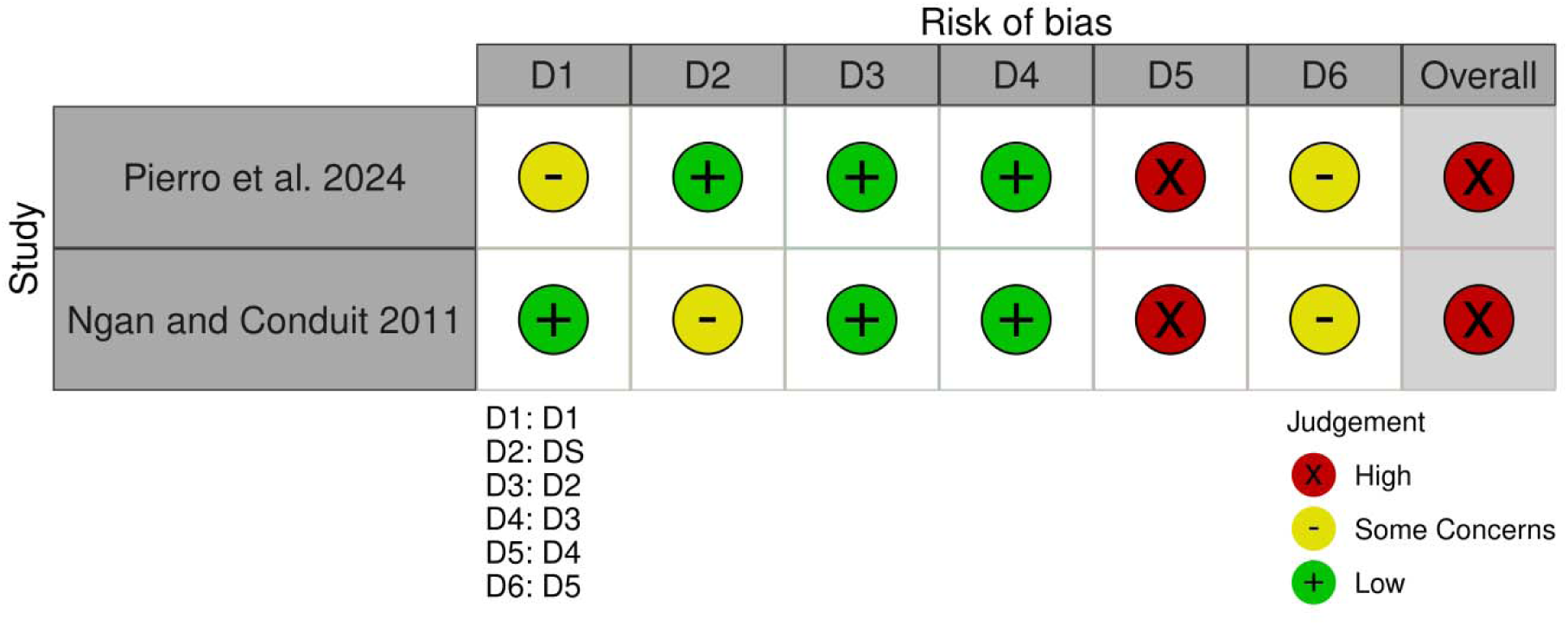
Risk of bias assessment crossover trial.

### Risk of Bias Assessment in Nonrandomized Controlled Trials

The risk of bias of nine included nonrandomized studies was evaluated and rated as the overall critical risk of bias in the majority of the studies. The overall ratings demonstrate that seven out of nine studies have a critical risk of bias: Catherine et al. (2021) [74]; Feyzabadi et al. (2014) [75]; Cases et al. (2011) [76]; Kim et al. (2024) [77]; Kimtata et al. (2023) [79]; Lemoine et al. (2019) [80] and Jaroenngarmsamer et al. (2021) [81]. The other two studies, Abdellah et al. (2020) [73] and Simone et al. (2024) [78], were rated as having a high risk of bias. No low or moderate overall risk of bias studies were found. The major reason for critical or serious risk of bias was due to Domain 1 (confounding), where eight studies were rated as a serious risk and Cases et al. (2011) [76] as a critical risk. Domain 6 (selection of participants) was also a problem, with eight out of nine studies rated as serious due to the selection of participants. Figure 4 shows risk of bias assessment for nonrandomized controlled trials

**Figure 4:**
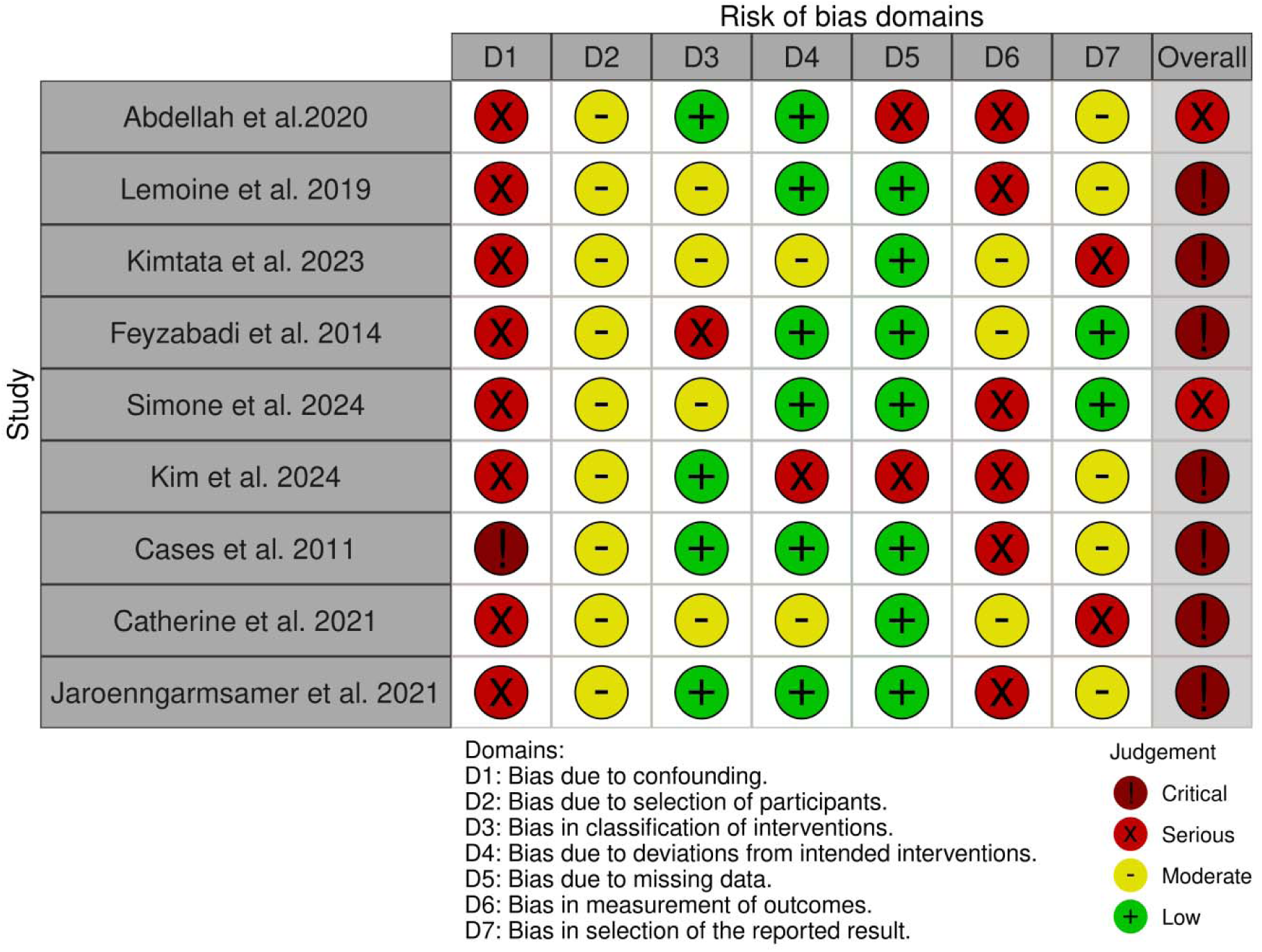
Risk of bias assessment for nonrandomized controlled trials.

## RESULTS OF INDIVIDUAL STUDIES

Table 3 presents the results of the continuous outcomes of all individual RCTs.

**Table 3:**
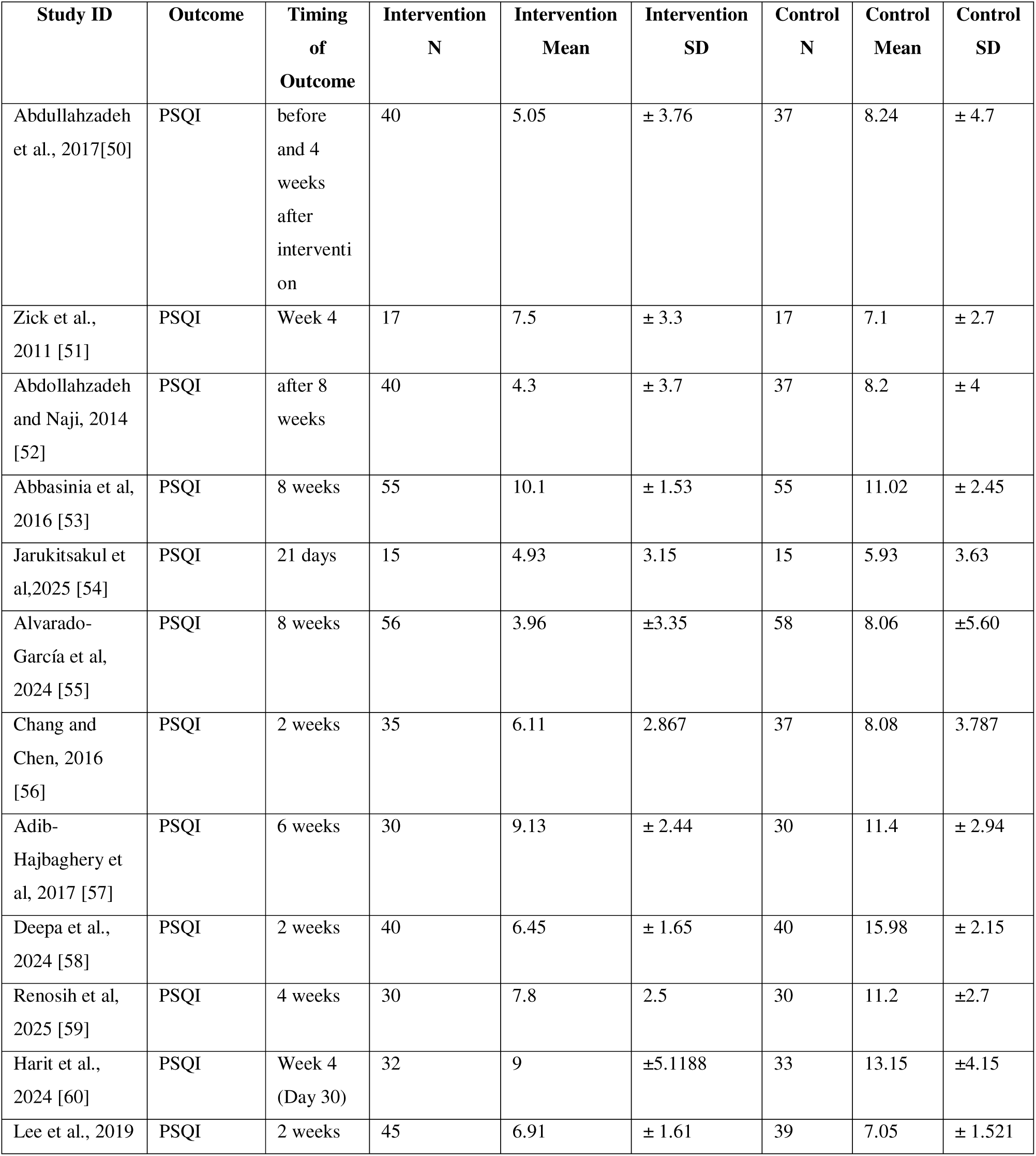
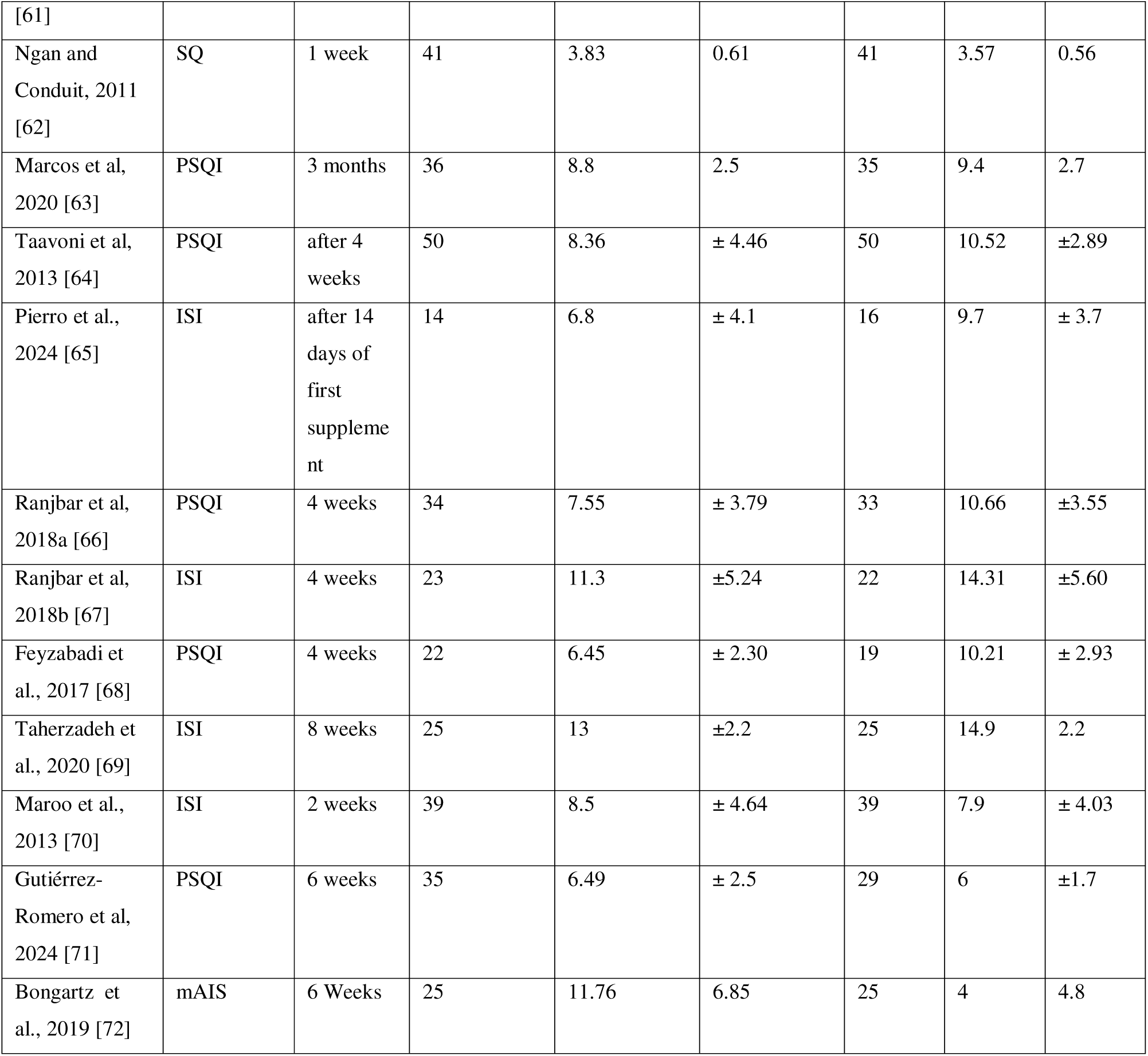
Results of continuous outcomes of individual RCTs.

**Table 4:**
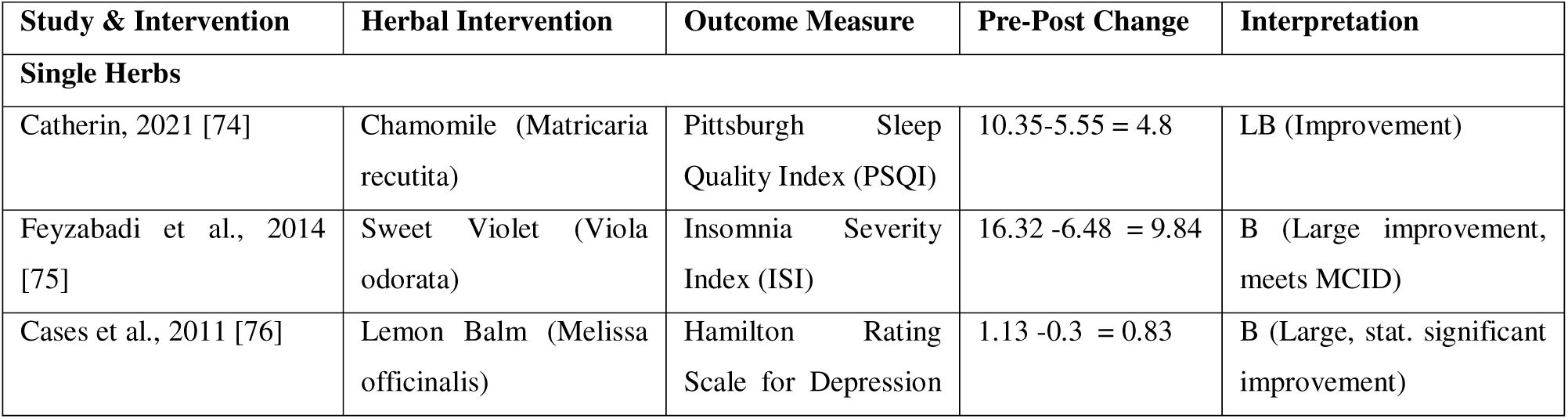
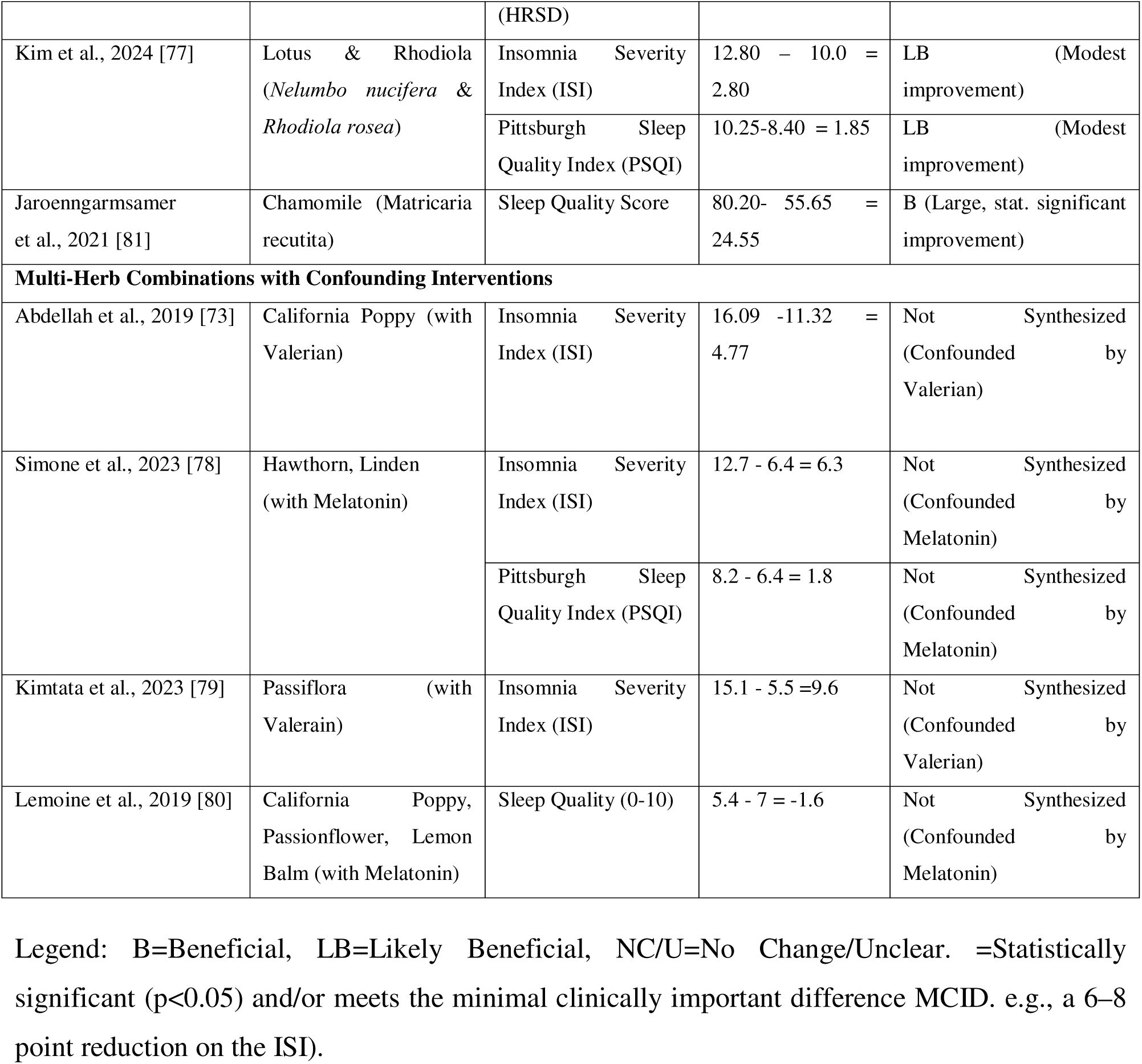
Results of the continuous outcomes of individual non-RCTs.

**Table 5:**
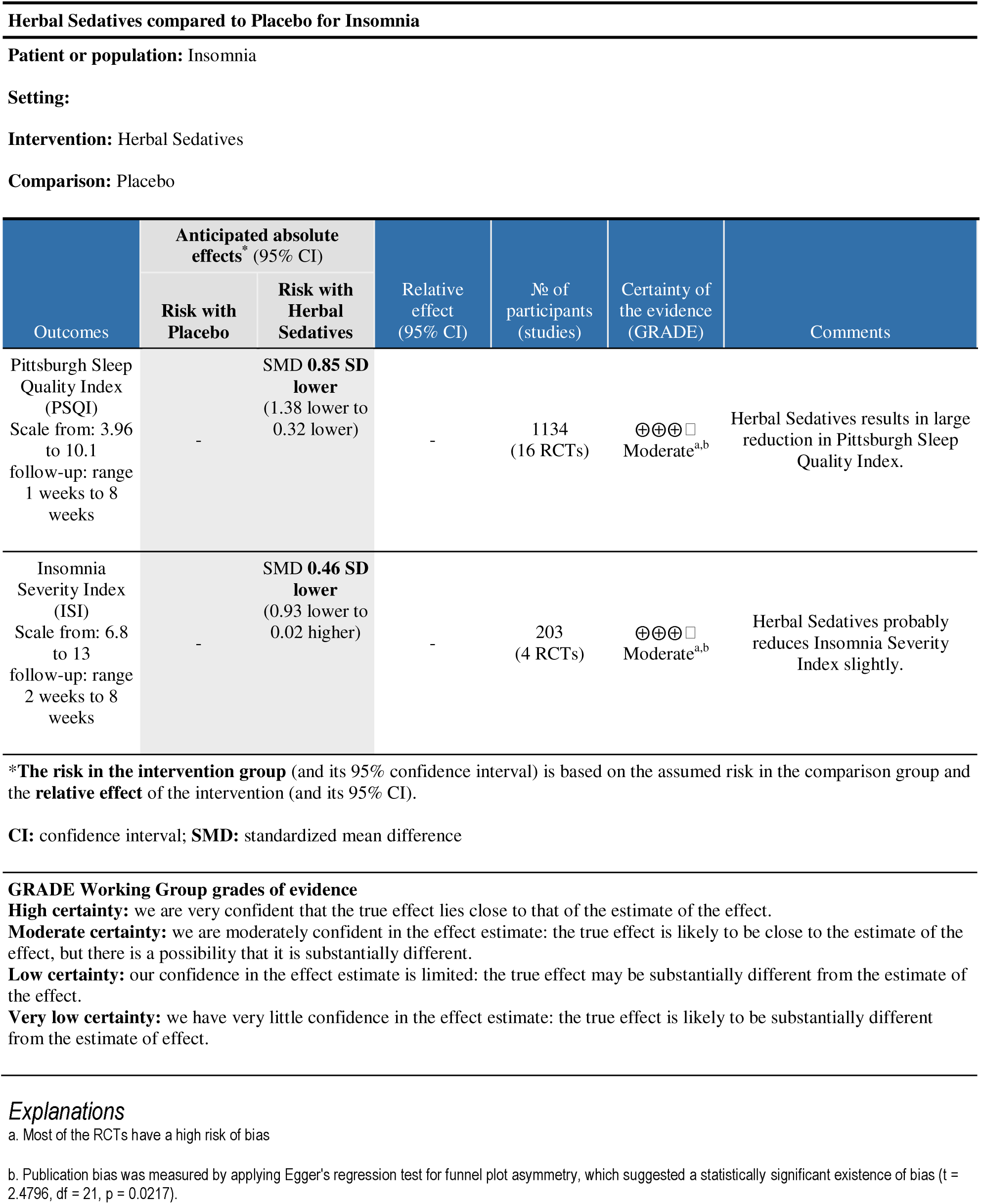
GRADE Summary of findings:

### Results of Syntheses

#### Narrative Synthesis

Nine studies with participants (n=369) were considered for narrative synthesis. The results are synthesized below, with a focus on prespecified single herb or multiple herb combination interventions.

### Synthesis by Intervention Group

#### 1. Single herbs

**Chamomile (Matricaria recutita):** Evidence from two studies (n=80) indicates that chamomile essential oil aromatherapy is beneficial for improving sleep quality. Jaroenngarmsamer et al. (2021) [74] reported a large, statistically significant improvement (p=0.022), and Catherin et al. (2021) [81] reported a clear positive change in the PSQI.

**Sweet Violet (Viola odorata):** One study (Feyzabadi et al., 2014 [75], n=50) provided evidence that intranasal sweet Violet oil is beneficial, demonstrating a large, clinically important reduction in ISI scores (−9.84 points).

**Lemon Balm (Melissa officinalis):** The evidence from Cases et al. (2011) [76] (n=20) is significant clinical improvement from the baseline score on the HRSD.

**Lotus & Rhodiola (*Nelumbo nucifera* & *Rhodiola rosea***): One study (Kim et al., 2024 [77], n=20) reported beneficial but modest improvements in both the ISI (2.80) and PSQI (1.85). The high attrition rate (35%) in this study lowers the confidence in this finding.

#### 2. Multi-Herb Combinations with Confounding Interventions

Two studies (Abdellah et al., 2019 [73] and Kimtata et al., 2023 [77], total n=136) investigated combinations that included a prespecified herb (California Poppy and Passionflower, respectively) but also included Valerian, which was not a prespecified intervention. While both studies reported positive outcomes, the effect of the prespecified herb cannot be isolated from that of Valerian. Therefore, these studies are notable but do not contribute to the synthesis of evidence for any single prespecified herb, as their results are confounded.

Two studies (Lemoine et al., 2019 [78] and Simone et al., 2023 [80]) (n=62) investigated combinations of prespecified herbs. Lemoine et al. (2019) [78], using a combination of California Poppy, Passionflower, and Lemon Balm, reported a statistically significant improvement in subjective sleep quality (p<0.001). Simone et al. (2023) [80], using a combination of Hawthorn and Linden, reported a clinically important reduction in ISI scores (−6.3 points). However, it is not possible to attribute the effects to any single herb within these combinations, and they do not contribute to the synthesis of evidence, and their results are confounded by melatonin.

### Meta-Analysis

We performed a meta-analysis of 23 randomized controlled trials (RCTs) that compared herbal sedatives with a placebo or control group. Three out of five outcomes were measured on reverse scales, and their effect sizes were inverted accordingly to ensure that a consistent movement of the effects was measured. A random-effects meta-analysis combining all 23 studies produced a standardized mean difference of −0.77 (95% CI [−1.14, - 0.40]), which was statistically significant (p=0.0001) and reflected the large pooled effects of herbal sedatives on sleep outcomes. Substantial and statistically significant heterogeneity was found among the studies (Q [df=22] = 149.17, p < 0.00001; I^2^ = 92%). Figure 5 shows Meta-Analysis of the effects of Herbal Sedatives on Sleep Outcomes

**Figure 5:**
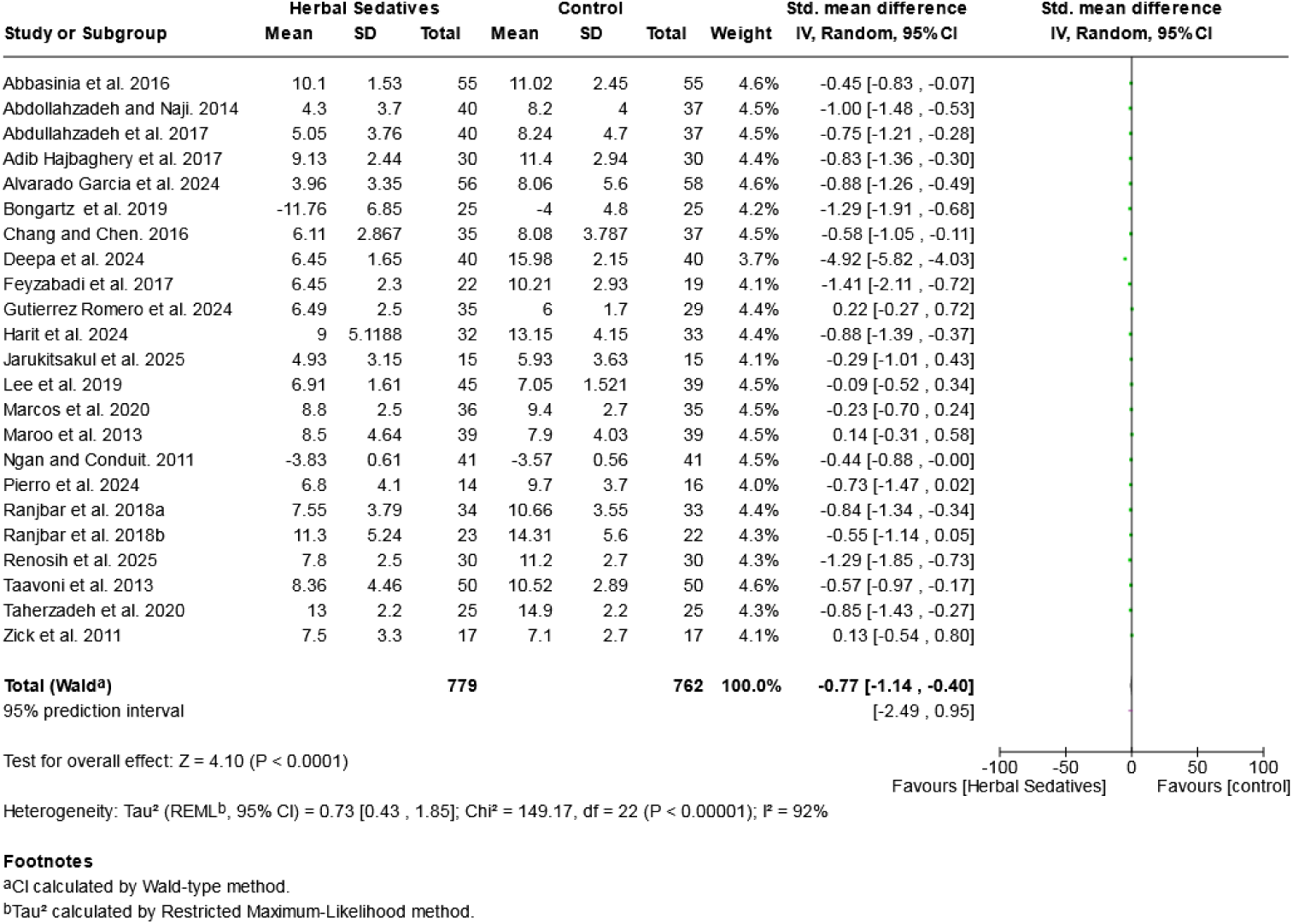
Meta-Analysis of the effects of Herbal Sedatives on Sleep Outcomes.

This forest plot illustrates the effect sizes of individual studies and the overall results of the meta-analysis. Each study is represented by a square (the standardized mean difference, SMD) and a horizontal line (the 95% confidence interval, CI). The size of each square reflects its contribution to the pooled result. The pooled SMD, calculated via a random-effects model, is shown as a diamond at the bottom. An SMD below zero favours the intervention, resulting in a reduction in outcome scores.

### Subgroup analysis

#### 1. Types of herbal sedatives

Subgroup analyses were conducted by type of herbal sedative in an attempt to investigate the potential sources of heterogeneity and to investigate whether particular herbal preparations might have differential effects.

For the 10 studies of chamomile, a significant improvement in sleep outcomes was found compared with the control (SMD= −1.06, 95% CI [−1.88, −0.24]; p= 0.01), even though significant heterogeneity was present (I² = 96%, χ² = 98.37, p < 0.00001).

For the four studies on Melissa officinalis, a significant beneficial effect was observed (SMD = −0.66, 95% CI −0.92 to −0.40; p < 0.00001), with no evidence of heterogeneity (I² = 0%, χ² = 0.84, p = 0.84).

For the four studies evaluating Passiflora, a small but statistically significant difference in favour of Passiflora was found (SMD (−0.39, 95% CI [−0.72, −0.07]; p = 0.02), with moderate heterogeneity (I² = 49%, χ² = 5.95, p = 0.11).

For the four studies in which herbal remedies were given in combination, the pooled effect was nonsignificant (SMD-0.42, 95% CI [−1.15, 0.30], p=0.25), and high heterogeneity predominated (I² = 87%, χ² = 21.47, p < 0.0001)).

The single study that assessed sweet violet extract revealed a large and statistically significant effect in favour of the intervention (SMD=-1.41 (95% CI [−2.11, −0.72]; p<0.0001).

A test for subgroup differences revealed no significant differences between groups of herbs (χ² = 8.38, df = 4, p = 0.08; I² = 52.2%), indicating that possible differences in the effect sizes of the different herbal types were not statistically significant. Figure 6 presents Subgroup analysis by type of intervention

**Figure 6:**
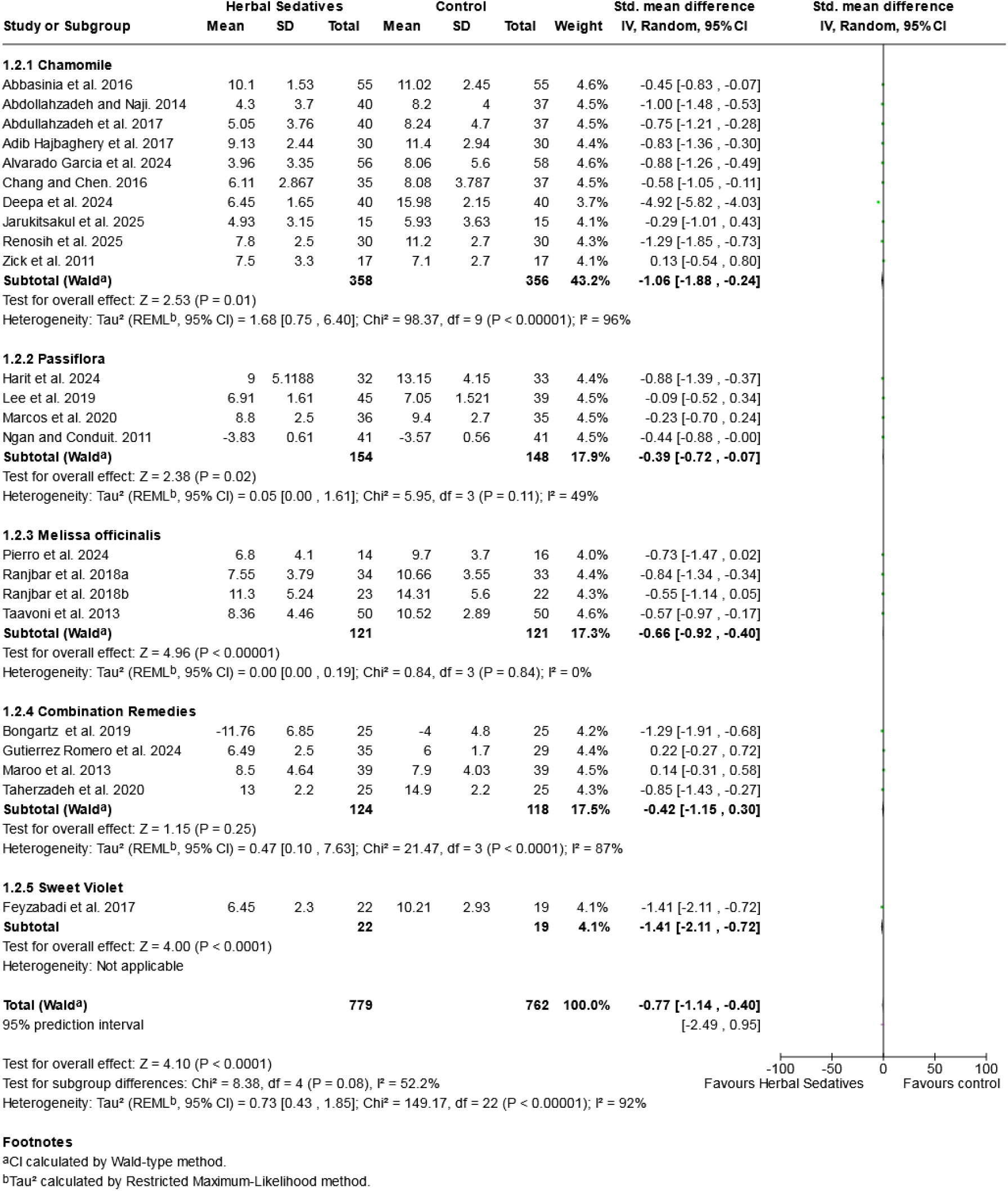
Subgroup analysis by type of intervention.

#### 2. Mode of administration

Subgroup analysis was also conducted for mode of administration to determine whether there was any difference in effectiveness between oral and intranasal delivery. For 19 RCTs that used oral administration, we found a statistically significant improvement in sleep outcomes compared with the control (SMD than −0.53, 95% CI [−0.73--0.34]; p < 0.00001), with a moderate pooled effect but substantial heterogeneity (I² = 65%, χ² = 50.12, p < 0.0001).

For the 4 studies that used intranasal administration, a greater overall effect was observed in favour of the intervention (SMD = −1.99, 95% CI [-3.88, −0.10]; p = 0.04), although very high heterogeneity was observed (I² = 97%, χ² = 69.64, p < 0.00001).

A test for subgroup differences revealed no statistically significant difference between the oral and intranasal routes (χ² = 2.26, df = 1, p = 0.13; I² = 55.8%). Figure 7 depicts subgroup analysi by mode of intervention.

**Figure 7:**
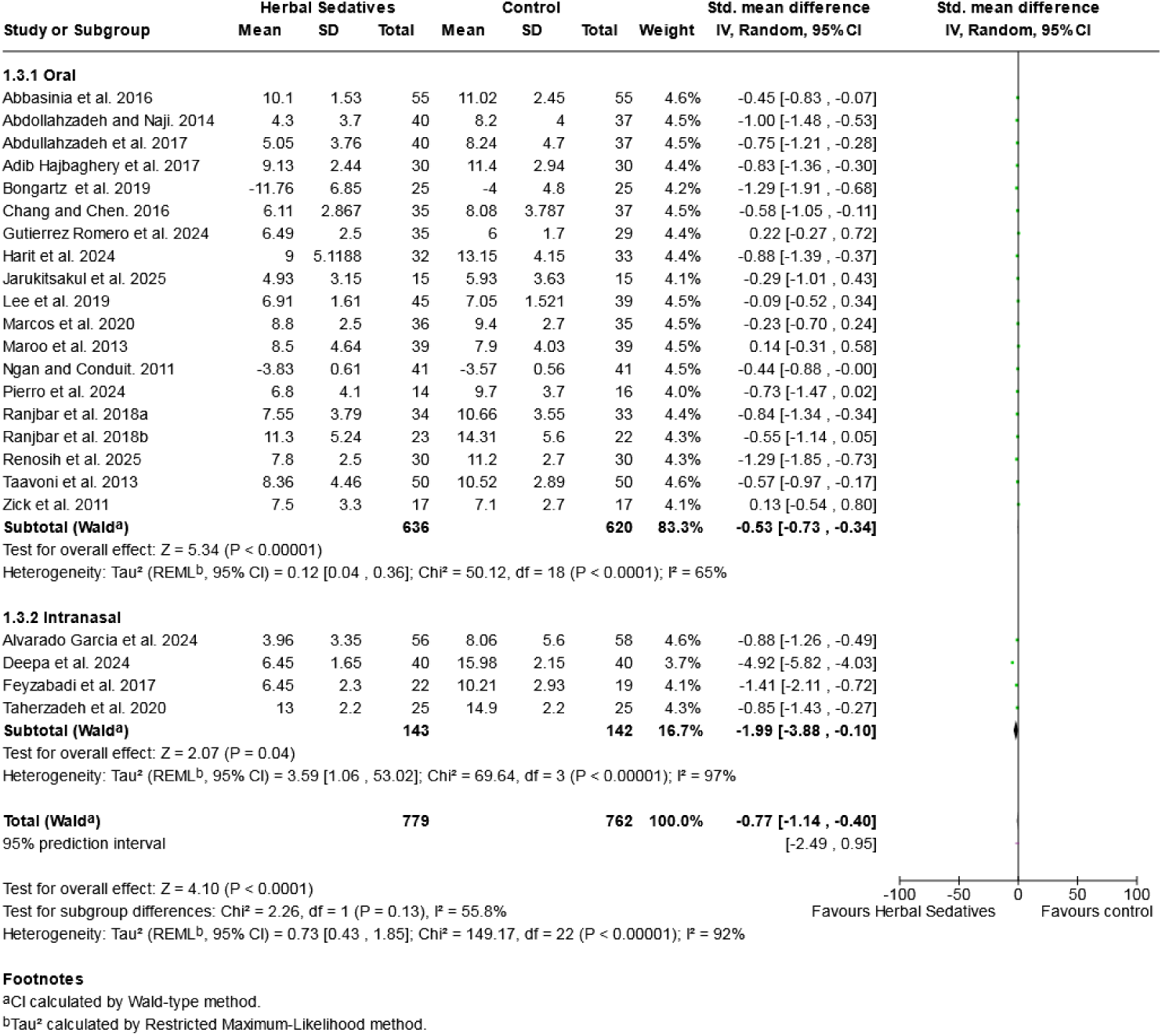
Subgroup analysis by mode of intervention.

### Sensitivity analysis

Sensitivity analyses were conducted to test the robustness and stability of the pooled effect estimate. The overall effect was statistically significant across a range of different analysis scenarios, suggesting that the beneficial effect was not caused by a single influential study. The pooled SMD was relatively stable and was uniformly in favour of intervention when individual studies were reviewed.

A sensitivity analysis was conducted to investigate the effects of combination remedies on the overall results. The overall standardized mean difference (SMD) was −0.77 (95% CI −1.14 to - 0.40) before the analysis. The exclusion of RCTs that used combination remedies and the restriction of study analysis to single herb interventions led to a larger effect size, SMD = −0.84 (95% CI −1.27, −0.42), suggesting a slightly greater degree of treatment effect at the same level of heterogeneity, i.e., I^2^=92%. The direction of the effect was consistent, indicating that the underlying beneficial effect of the outcomes in the primary meta-analysis was robust. Figure 8 shows sensitivity analysis for single herb versus combination remedies

**Figure 8:**
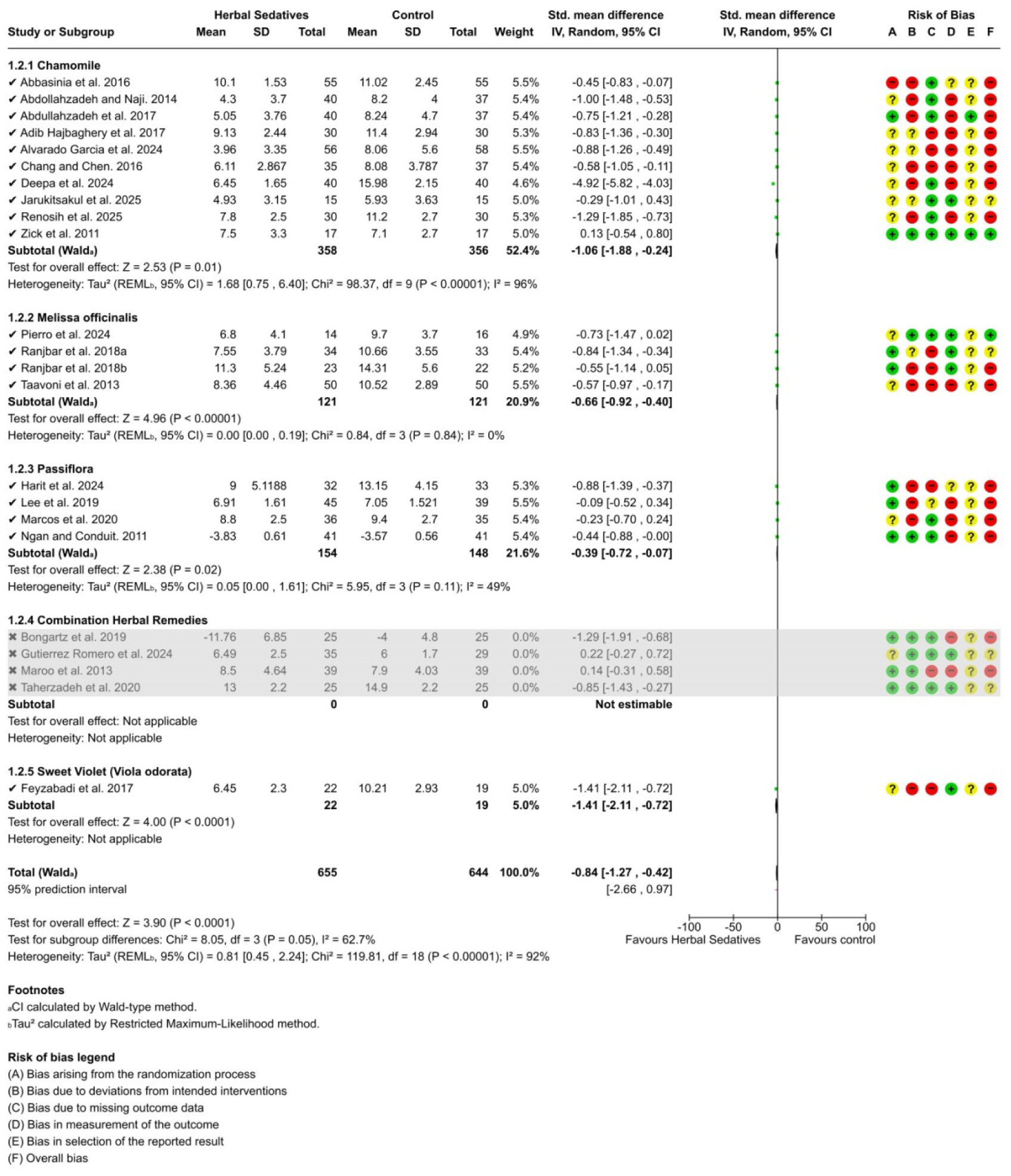
Sensitivity analysis for single herb versus combination remedies.

To further explore the high degree of heterogeneity found in the primary analysis (I^2^ = 92%), a sensitivity analysis was also conducted for the intervention mode of administration. Exclusion of studies using intranasal administration led to a decreased pooled effect size (SMD = −0.53; 95% CI −0.73-**-**0.34) and reduced heterogeneity to I^2^ = 65%. These results indicate that intranasal interventions were responsible for the greater overall effect size and greater between-study heterogeneity; however, the direction and statistical significance of the effect were the same across the studies. Figure 9 presents sensitivity analysis for mode of administration

**Figure 9:**
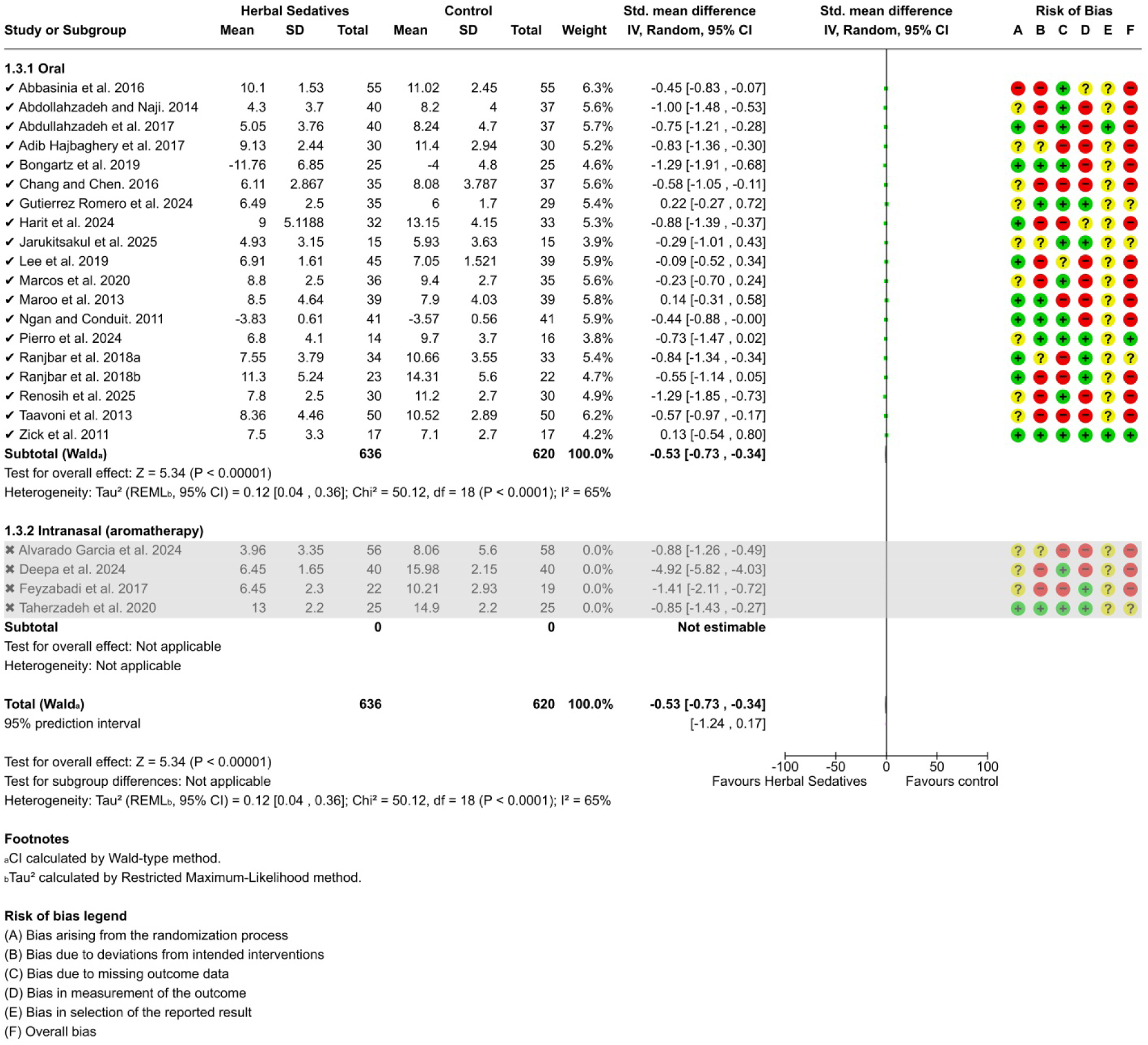
Sensitivity analysis for mode of administration.

Overall, the findings from sensitivity analyses suggest that the observed beneficial effect of herbal sedatives on sleep outcomes is relatively robust, although the magnitude of the effect may depend on the type of intervention and mode of administration.

### Publication Bias

Publication bias was measured via Egger’s regression test for funnel plot asymmetry, which suggested a statistically significant existence of bias (t = 2.4796, df = 21, p = 0.0217). The asymmetry observed in the funnel plot (Figure 10) provides visual confirmation of potential publication bias.

**Figure 10:**
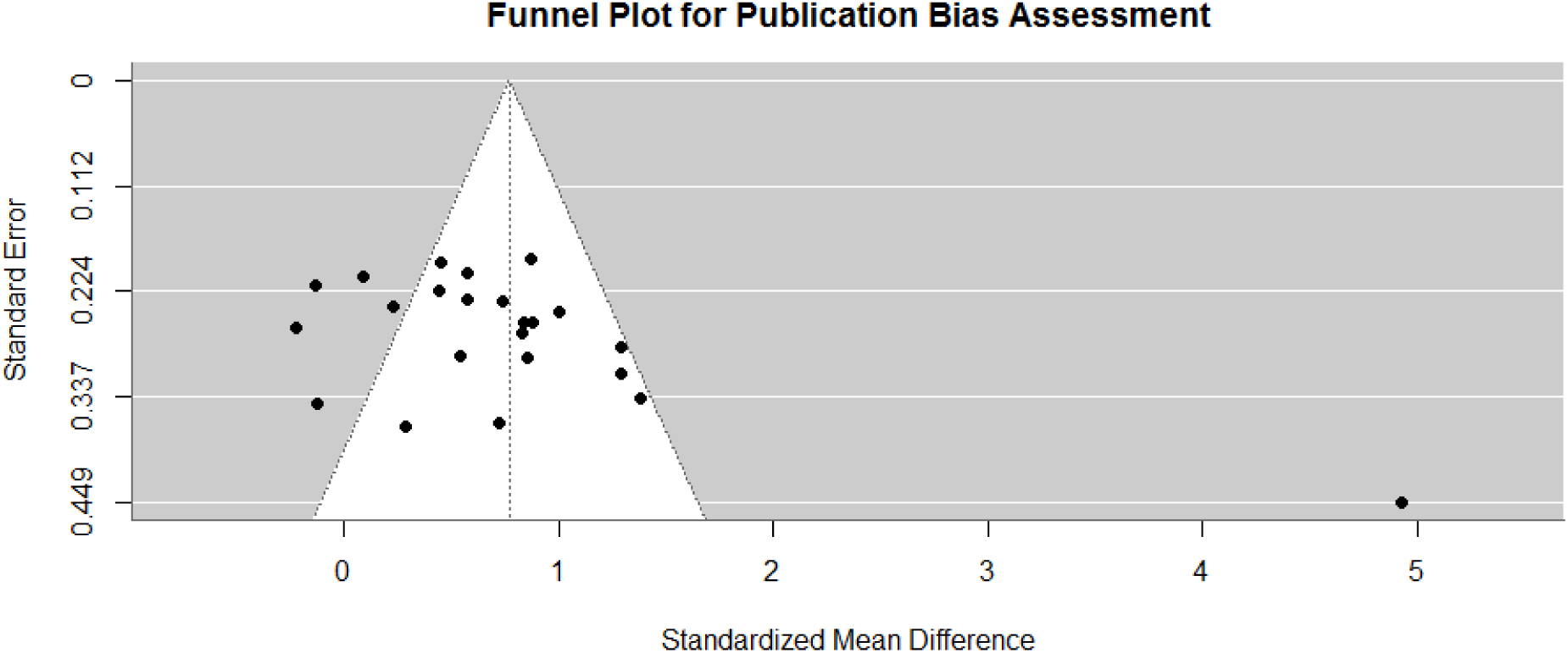
Funnel plot for publication bias assessment.

**Figure 11:**
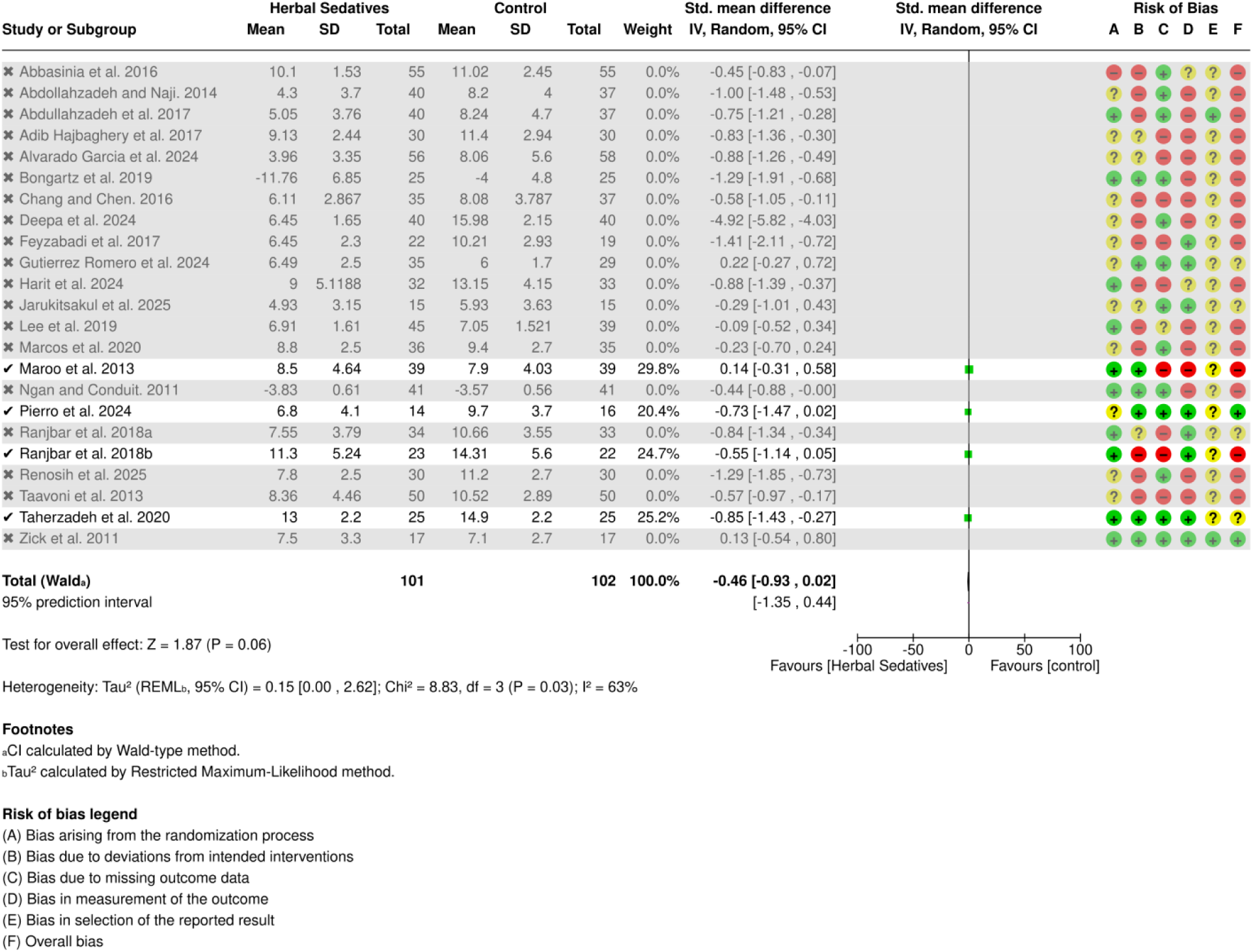
Forest Plot based on studies utilizing ISI Outcome Measures as cited Certainty of Evidence Table.

**Figure 12:**
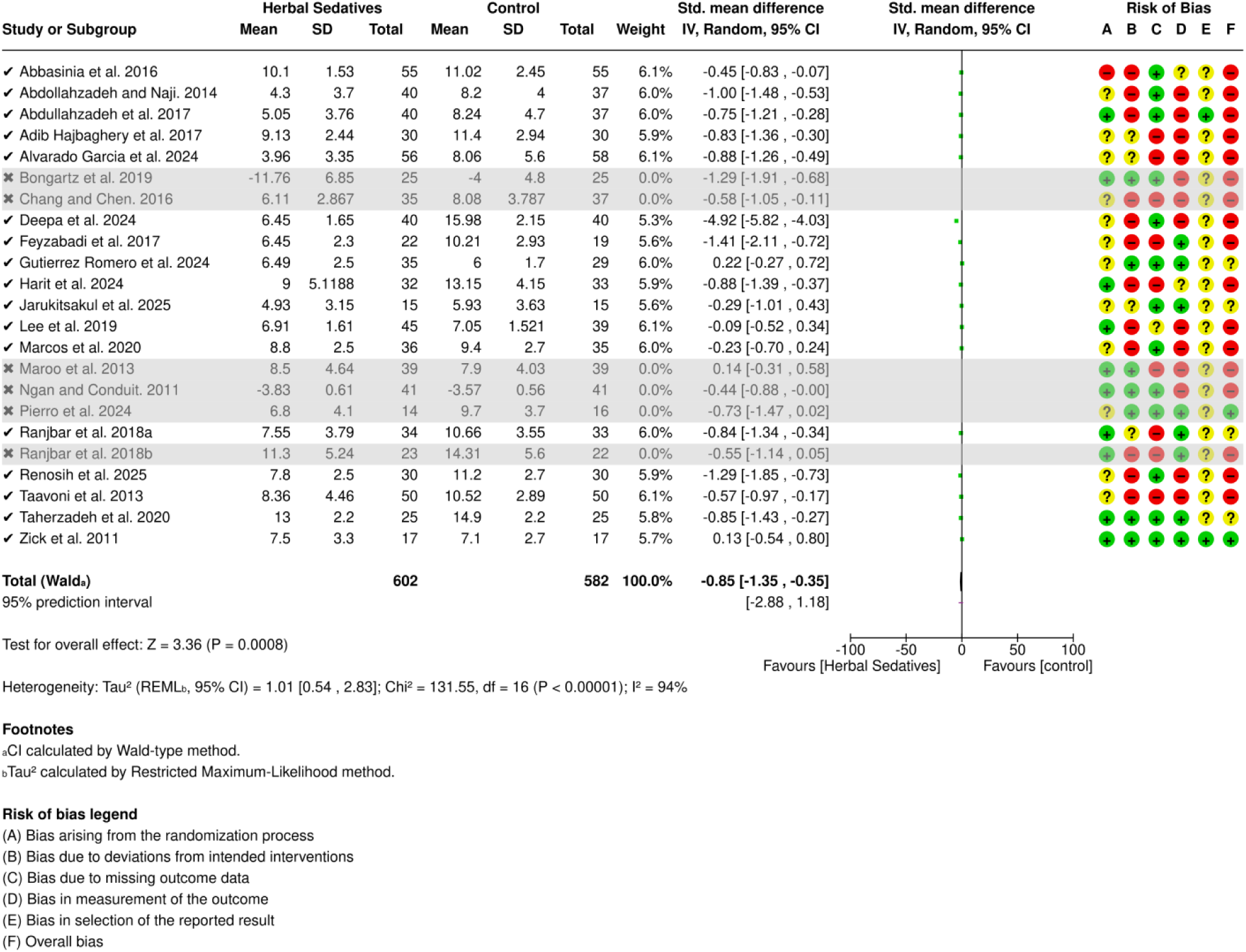
Forest Plot based on studies utilizing PSQI Outcome Measures Certainty of Evidence Table.

Figure 10 Funnel plot assessing the risk of publication bias by plotting each study’s effect (SMD) against its precision (standard error). The vertical dashed line represents the overall pooled effect, whereas the diagonal dashed lines represent the expected 95% confidence interval for a given standard error in the absence of bias. The observed asymmetry in the plot suggests that smaller studies with null or negative results may be missing from the analysis.

### Certainty of Evidence

The overall certainty of the body of evidence was judged by two reviewers (MAP and MDK) working separately. To perform this evaluation, we used the GRADE system implemented in the GRADEpro Guideline Development Tool [49]. The step-by-step assessment was performed for all five core domains for two major outcome measures, i.e., the Pittsburgh Sleep Quality Index (PSQI) and **the** Insomnia Severity Index (ISI).

Sixteen randomized controlled trials including 1134 participants reported sleep quality measured by the Pittsburgh Sleep Quality Index. Compared with placebo, herbal sedatives resulted in a large reduction in the Pittsburgh Sleep Quality Index (SMD −0.85* SD lower, 95% CI 1.38 lower to 0.32), but the evidence has a moderate level of certainty. The evidence was downgraded from high to moderate certainty, as most of the RCTs had a high risk of bias and potential publication bias. Four randomized controlled trials including 203 participants whose sleep outcomes were measured by the Insomnia Severity Index (ISI) revealed that, compared with placebo, herbal sedatives slightly reduced the insomnia severity index (SMD −0.46* SD lower, 95% CI 0.93 lower to 0.02 higher), but the certainty of evidence was rated as moderate due to a high risk of bias and potential publication bias across the broader body of evidence.

*\*The forest plots for these SMD scores are attached as supplementary files*.

## Discussion

This systematic review and meta-analysis synthesized evidence from 23 randomized controlled trials evaluating herbal interventions for insomnia. The pooled analysis demonstrated a moderate overall treatment effect (SMD = −0.77) favouring herbal interventions compared with placebo or the control. Sensitivity analysis further indicated that single-herb interventions produced a slightly greater pooled effect (SMD = −0.84) than combination herbal remedies did (SMD = - 0.77). Chamomile and Melissa officinalis (lemon balm) are the most frequently investigated botanicals and have consistently improved sleep quality across several trials. This review provides a more detailed synthesis of the therapeutic potential of botanical sleep aids than previous narrative or condition-specific reviews do by quantitatively comparing individual and combined herbal preparations. The present meta-analysis extends this literature by synthesizing evidence across a broader range of herbal interventions and exploring differences between single-herb formulations and combination formulations through subgroup analysis.

The strongest pooled effect in our analysis was observed for chamomile, which confirmed a large effect size relative to other herbal interventions. This finding is broadly consistent with the emerging clinical literature. For example, a recent randomized controlled trial compared the effects of chamomile and Passiflora on sleep quality among 90 insomnia patients aged 25--45 years and reported that chamomile significantly improved insomnia and daytime functioning compared with Passiflora [82]. Similarly, a systematic review and meta-analysis of clinical trials examined the effects of chamomile on sleep outcomes and reported a significant reduction in the Pittsburgh Sleep Quality Index (PSQI) score (WMD: −1.88, 95% CI) [83]. Chamomile, a member of the Asteraceae family, has long been used in traditional medicine for insomnia, fever, muscular spasms, menstrual problems, ulcers, gastrointestinal issues and rheumatic pain [84]. Its therapeutic effects are thought to be mediated primarily through bioactive compounds such as apigenin, which acts as an antagonist of gamma aminobutyric acid (GABA) receptors and exhibits anxiolytic and sedative properties [85]. Despite its widespread traditional use, authoritative reviews, such as those by the National Centre for Complementary and Integrative Health (NCCIH), highlight that robust clinical evidence supporting chamomile efficacy for insomnia remains limited [86, 87]. Experimental studies have provided mixed support for the hypnotic properties of chamomile. For example, an earlier in vivo study investigating the hypnotic effects of chamomile and Passiflora extracts in animal models revealed significant shortening of sleep latency in chamomile but no effects on sleep latency or delta activity during non–REM (rapid eye movement) with Passiflora [88]. Similarly, a systematic review examining the therapeutic effects of chamomile, which included 69 studies from 1990--2016, revealed its beneficial effects as an antioxidant and antidepressant and was helpful in treating osteoarthritis and ulcerative colitis but did not report its benefits for insomnia [89]. Compared with those of conventional pharmacological treatments, the magnitude of chamomile effects is notable. Benzodiazepine receptor agonists and z-drugs have an SMD between 0.57 and 0.78 and are related to well-documented profiles of adverse effects (dependency and next-day sedation) [90]. The similar effect size compared with that of chamomile, combined with a positive safety profile, therefore represents a clinically useful signal; however, this finding needs to be confirmed by greater certainty before it can be implemented in clinical practice.

Our subgroup analysis by intervention type revealed that Melissa officinalis (lemon balm) had the second largest effect size, i.e., −0.66. This finding aligns with contemporary clinical evidence suggesting beneficial effects of lemon balm on sleep and anxiety. A narrative review investigated the effects of Melissa officinalis on sleep quality and reported that lemon balm extracts reduce sleep disorders in cardiac patients [91]. In addition, a double-blind randomized placebo-controlled trial examining the efficacy of lemon balm for anxiety and insomnia in coronary artery bypass surgery patients reported a reduction in anxiety scores, i.e., 10.18 in the control group and 7.15 in the lemon balm group, p = 0.001, and substantial improvements in sleep quality score, i.e., 14.40, compared with the control, i.e., 7.52, p < 0.001, in the lemon balm [92]. The pharmacological basis for this moderate effect size is biologically plausible, as the lemon balm contains several active compounds, such as rosmarinic acid, which inhibits gamma aminbutyric acid transaminase (GABA-T) [93]. This inhibition results in increased synaptic availability of neurotransmitters. Gabapentin mimics the anxiolytic effect instead of the direct hypnotic action to promote sleep [93]. This distinction is clinically important because, unlike direct hypnotic agents that activate gamma-aminobutyric acid (GABA-A) receptors, this mechanism likely produces a milder sedative profile and may therefore reduce the risks associated with stronger sedative medications.

In contrast, for chamomile and lemon balm, the subgroup analysis of Passiflora yielded a small pooled effect of 0.39. However, both experimental and clinical evidence suggests that Passiflora may still exert beneficial effects on sleep physiology. Animal studies provide some support for this hypothesis. For example, Guerrero and Medina (2017) examined the effect of Passiflora incarnata on Wistar rats via electroencephalography (EEG) recordings and reported a significant improvement in total sleep time (p<0.05) [94]. Similarly, Kim et al. (2019) investigated the sleep-inducing effect of Passiflora incarnata in rodents and reported that Passiflora extracts influenced GABA receptor activity and significantly increased the resting time following the oral administration of 250–500 mg of Passiflora [95]. Recent human studies also support our findings. A double-blind randomized controlled trial investigated the effects of Passiflora on polysomnographic sleep parameters. A total of 110 adult participants received Passiflora for 2 weeks, and the total sleep time (TST) was significantly greater in the Passiflora group (23.05 ± 54.26) than in the placebo group (−0.16 ± 53.12 [96]. A retrospective observational study by La Tempa et al. (2025) explored the use of Passiflora incarnata for insomnia among children and adolescents and recruited 94 patients with anorexia nervosa who were administered 200 mg of Passiflora incarnate with selective serotonin reuptake inhibitors (SSRIs) and reported that 45.4% of patients reported improvements in insomnia [97]. Although these findings are encouraging, further high-quality trials are needed to clarify the magnitude and consistency of the therapeutic effects of Passiflora.

Our meta-analysis included a single RCT by Feyzabadi et al. (2017) [68] examining violet oil, which reported a relatively large effect size (SMD = −1.41). However, because this estimate was derived from a single study with a high risk of bias, its reliability is limited. Another trial by Taherzadeh et al. (2020) [69] evaluating violet oil in combination with saffron and lettuce seeds reported a somewhat smaller effect size, i.e., an SMD of 0.85, indicating its beneficial effects both as a stand-alone intervention and as part of multicomponent formulations. Nonetheless, the available evidence remains sparse. A previous systematic review and meta-analysis assessing the effect of Violet extract on insomnia included five clinical trials that reported improvements in total PSQI scores (MD, −4.67; P = 0.0002), sleep duration scores (MD, −0.77; P < 0.00001), and ISI scores (MD, −6.30; P = 0.009) in the Violet extract group compared with the placebo group [98]. These findings suggest promising effects, but additional high-quality RCTs are needed to confirm their efficacy.

In addition to the meta-analysis of randomized trials, our review also examined evidence from nonrandomized studies through narrative synthesis. This included *Rhodiola rosea* and *Nelumbo nucifera*, both of which demonstrated moderate improvements in sleep-related outcomes in preliminary research. Experimental studies indicate that *Nelumbo nucifera* (lotus) extract may possess sleep-promoting properties; for example, a recent study in an animal model investigated the sleep-promoting activity of *Nelumbo nucifera* (lotus) in mice and reported that 50 mg of lotus extract improved sleep duration by 24% compared with that of the control when it was orally administered [99]. Similarly, a study in humans investigated the effects of *Rhodiola rosea* on stress among ninety patients with obstructive sleep apnea and revealed that Rhodiola may reduce stress by inhibiting oxygen free radicals [100]. Traditionally, *R. rosea* has been used to treat depression, anxiety, and fatigue and to improve the ability to concentrate [101, 102]. Its adaptogenic properties are believed to involve modulation of the hypothalamic pituitary adrenal (HPA) axis, and cortisol regulation is a logical yet unconfirmed mechanism for improving sleep as well in stress-related insomnia [103]. However, clinical evidence evaluating Rhodiola specifically for insomnia remains extremely limited, highlighting an important research gap.

Our narrative synthesis also included one nonrandomized control trial of Eschscholtzia californica, which was administered in combination with Chamomile and Valeriana officinalis. This herb contains alkaloids with documented sedative, anxiolytic, and analgesic effects [104]. Eschscholtzia californica is also utilized as a homeopathic remedy for insomnia in its crude form. A small comparative study with 30 insomnia patients revealed that Eschscholtzia californica mother tinctures produced greater improvements in sleep symptoms than did Passiflora mother tinctures [105]. While these findings suggest possible therapeutic potential, the limited number of trials and methodological weaknesses prevent definitive conclusions regarding efficacy.

Another botanical of interest was Crataegus (Hawthorn), which appeared only in combination formulations with Linden, vitamin B1, and melatonin. Because of these confounding ingredients, it was not included in the narrative synthesis. A recent randomized double-blind controlled trial examining the efficacy of Hawthorn fruit extract on blood pressure and quality of sleep among 60 patients with hypertension and sleep disorders reported significant improvements in the Hawthorn group compared with the control group (P = 0.001) [106], but our review remains inconclusive with regard to this remedy owing to the coadministration of other sleep-modulating agents.

Several RCTs and non-RCTs included in the meta-analysis investigated combination herbal remedies. These formulations produced a pooled effect size of −0.42, which was smaller than the overall pooled estimate. When combination interventions were excluded from the sensitivity analyses, the pooled effect increased from 0.77 to 0.84, suggesting that single-herb formulations may produce more consistent therapeutic effects. This finding challenges the common assumption within phytotherapy that multiherb formulations necessarily produce synergistic benefits. One plausible explanation may involve pharmacokinetic interactions or receptor-level competition among bioactive constituents, although this hypothesis requires further pharmacological investigation; concurrent administration of multiple acts of both the GABAergic and serotonergic families may lead to receptor saturation, metabolic competition at CYP450 family pathways, or partial antagonism at shared receptor locations [107]. Another possibility is that combining multiple botanical ingredients may dilute the concentration of key bioactive compounds from individual plants, potentially reducing the effective pharmacological dose delivered by each component. Because the included combination trials differed substantially in composition, dosage, and standardization, definitive conclusions about synergistic interactions remain difficult.

### Limitations of Evidence

The clinical implications of the pooled SMD of 0.77 to 0.84 need to be viewed in light of the significant statistical and clinical heterogeneity across the included trials. Differences in participant populations, intervention formulations, outcome measures, and study designs likely contributed to variability in treatment effects. Moderate-to-high heterogeneity (I²) suggests that the pooled estimate may reflect substantial underlying differences among studies, potentially limiting generalizability [108].

Using the GRADE framework, the overall certainty of evidence was rated as moderate, primarily because of methodological weaknesses, high risk of bias in most included RCTs, significant clinical heterogeneity, and the lack of large-sized, preregistered, multicenter trials [45, 48]. The evidence should be interpreted with the consideration that most of the included RCTs (except four) were at high risk of bias, which severely limits confidence in the pooled effect estimate.

A further challenge in phytomedicine research involves the translational gap between preclinical and clinical evidence. Animal studies often employ nonstandardized extract concentrations and drug-induced sleep models that may not accurately reflect human insomnia physiology. Differences in sleep architecture between rodents and humans make cross-species extrapolation methodologically unreliable [109]. Despite the widespread use of these complementary therapies for insomnia, there is an absence of clinical practice guidelines [110, 111]. The lack of these guidelines is a systemic failure to invest in rigorous clinical research on low-cost and low-risk interventions. This evidence practice gap is especially significant in low-resource settings where herbal sedatives are easily available options [112].

There are limitations at the level of individual herbs. For example, observational studies evaluating *Nelumbo nucifera* and *Rhodiola rosea* are the most important limitations. They provide only preliminary evidence and cannot establish causal relationships because of potential confounding factors and a lack of control groups [113]. The evidence base for Eschscholzia californica is similarly limited owing to evidence from a nonrandomized controlled trial that corresponds to the Oxford CEBM Level 3 evidence, which is susceptible to allocation bias, performance bias and confounding factors [114]. Evidence limitations also exist at the level of combination remedies where the observed therapeutic effect cannot be disaggregated for any single constituent, thus making the results noninterpretable. For hawthorn (Crataegus), the absence of standalone RCTs for insomnia prevents any definitive conclusions regarding efficacy, and it cannot be recommended as a hypnotic agent.

### Limitations of the review process

This systematic review also has several methodological limitations. First, combination remedies were excluded from the sensitivity analysis, which may have inadvertently removed trials with a higher risk of bias, potentially influencing the observed pooled effect size. Second, identifying certain single herbal interventions, such as sweet violet, *Rhodiola rosea*, Eschscholtzia californica and Crataegus, has proven challenging. Despite extensive database and trial register searches, we only found one RCT by Feyzabadi et al., 2017 [68] examining the use of Violet oil for insomnia that met the inclusion criteria; therefore, it could not be included in subgroup analyses. Similarly, Eschscholtzia californica appeared only in nonrandomized trials or combination products. We conducted a single observational study by Kim et al. (2024) [77] on *Rhodiola rosea* and *Nelumbo nucifera* and a single nonrandomized control trial by Abdellah et al. (2019) [73], examining Eschscholtzia californica in combination with chamomile and Valeriana officinalis or melatonin, vitamin B6, passionflower extract (150 mg), and lemon balm extract (240 mg) by Lemoine et al. (2019) [80]. We included Lemoine et al.’s (2019) [80] study in the narrative synthesis, as it contained three of our required interventions, i.e., Eschscholtzia californica, Passiflora and Lemon balm. Similarly, we could not find a single RCT or non-RCT for Hawthorn (Crataegus). The only study involving Crataegus, i.e., Simone et al. (2023) [78], used a multiingredient formulation including melatonin and vitamin B1, which was not included in the narrative synthesis because it prevents reliable interpretation of Hawthorn’s independent effects.

The narrative synthesis is limited by the primary studies available. The reliance on vote counting does not provide a pooled effect size. The most significant limitation is the paucity of studies providing direct, unconfounded evidence for each individual herb. For several herbs (Passionflower, Hawthorn, California Poppy), the only available evidence comes from multiple herb combinations, preventing definitive conclusions about their individual efficacy. The positive findings from confounded studies (involving Valerian) highlight a gap in the literature for pure investigations of prespecified herbs.

### Implications for Practice, Policy and Future Research

Despite these limitations, the relatively large pooled effect size observed in this review highlights the potential therapeutic relevance of certain herbal sedatives, particularly chamomile and lemon balm. The large effect size of chamomile and favourable safety profile are clinically meaningful signals for addressing critical issues in acute care and require further investigation [90]. These interventions may represent accessible and low-risk alternatives for individuals seeking nonpharmacological approaches to insomnia management. However, clinicians should interpret these findings cautiously until they are supported by larger, high-quality trials.

Future trials are needed to establish the efficacy of Melissa officinalis in primary and secondary insomnia subtypes. Additional research is also needed to clarify potential herb–drug interactions, particularly those involving serotonergic pathways. The interaction of herbal products with serotonergic pathways should receive pharmacovigilance attention because the risk posed by serotonin syndrome in combination with selective serotonin-uptake inhibitors (SSRIs) has not been systematically investigated [115]. Pharmacovigilance studies will therefore be essential to ensure safe clinical implementation.

Major evidence gaps remain for several commonly used botanicals. For example, the complete paucity of RCT evidence on the efficacy of *Rhodiola rosea*, Eschscholtzia Californica and Hawthorn for insomnia highlights the urgent need for future research. In particular, these sleep formulations are widely available worldwide. A substantial evidence gap is noted for Crataegus, and future studies are obligated to fill this gap through studies on diverse patient populations for primary insomnia as defined via validated diagnostic criteria (e.g., DSM-5).

The nonanticipated result of individual interventions being superior to combination remedies has significant clinical implications: clinicians should not assume that products with combinations of herbal constituents are better than single herb preparations in the absence of direct, randomized controlled trial (RCT) evidence; regulatory authorities may require that products with multiple ingredients be tested before they are allowed to bring multiple-ingredient sleep formulations to market [116]. Future research should prioritize comparisons of standardized single herb and combination preparations to elucidate optimal formulation strategies.

Future trials must conform to the CONSORT 2010 extension for herbal medicines, include detailed reporting of extract standardization, dosage, and treatment duration and be registered on ClinicalTrials.gov, including both objective sleep results (polysomnography) and validated subjective instruments (e.g., PSQI, ISI), and the use of active comparators as well as placebo as an option for establishing clinical equivalence thresholds. [117]. Without these advances in methodology, even a large pooled effect size should not be considered definitive enough evidence on which to base clinical guidelines or recommendations for prescribing regimens.

### Conclusion

In conclusion, this systematic review and meta-analysis provides preliminary evidence that several herbal sedatives may improve insomnia-related outcomes, with chamomile and Melissa officinalis demonstrating the most consistent therapeutic signals. However, the strength of this evidence remains limited by methodological weaknesses, a high risk of bias, and substantial heterogeneity across studies. Nevertheless, the biological plausibility of these herbal sedatives cannot be ruled out through mechanisms such as GABAergic modulation, anti-inflammatory activity, oxidative stress reduction, and circadian regulation [118]. These mechanisms must be confirmed through rigorous clinical research. Future studies demand standardized extracts, larger multicentre RCTs, and extended follow-up periods to establish their safety and efficacy. The statistically large pooled effect of the herbal sedatives must be interpreted with caution due to the overall low certainty and heterogeneity of evidence. The current evidence is not convincing enough to support whether these herbal sedatives are effective interventions for insomnia. Until such evidence becomes available, the use of herbal sedatives for insomnia should be considered prudently.

## List of abbreviations

**Title page and background:**

- AASM - American Academy of Sleep Medicine
- CBT-I - Cognitive Behavioral Therapy for Insomnia
- CAM - Complementary and alternative medicine
- FDA - Food and Drug Administration
- PSQI - Pittsburgh Sleep Quality Index (mentioned later but relevant)

**Methods:**

- PROSPERO - International Prospective Register of Systematic Reviews
- PRISMA - Preferred Reporting Items for Systematic Reviews and Meta-Analyses
- PICO - Population, Intervention, Comparator, Outcome
- CBT - Cognitive Behavioral Therapy
- CBT-I Cognitive behavioral therapy for insomnia
- ISI - Insomnia Severity Index
- ESS - Epworth Sleepiness Scale
- PSQI - Pittsburgh Sleep Quality Index
- WHO ICTRP - World Health Organization International Clinical Trials Registry Platform
- OTAD - Open Thesis and Dissertations
- RoB 2.0 - Risk of bias tool 2.0
- ROBINS-I - Risk of Bias in Nonrandomized Studies of Interventions
- SWiM - Synthesis Without Meta-analysis
- MCID - Minimal clinically important difference
- GRADE - Grading of recommendations, assessment, development, and evaluations
- SMD - Standardized mean difference
- CI - Confidence interval
- RevMan - Review Manager software

**Results:**

- mAIS - modified Athens Insomnia Scale
- SQ - Sleep quality
- RCT - Randomized Controlled Trial

**Discussion:**

- NCCIH - National Centre for Complementary and Integrative Health
- REM - Rapid Eye Movement
- GABA - Gamma Aminobutyric Acid
- GABA-A - Gamma aminobutyric acid type A receptor
- GABA-T - Gamma Aminobutyric Acid Transaminase
- TST - total sleep time
- SSRIs - Selective serotonin reuptake inhibitors
- HPA - Hypothalamic Pituitary Adrenal
- CYP450 - Cytochrome P450
- DSM-5 - Diagnostic and Statistical Manual of Mental Disorders, Fifth Edition
- CONSORT - Consolidated Standards of Reporting Trials
- HRSD - Hamilton Rating Scale for Depression

**Appearing in references and throughout:**

- DF - degrees of freedom
- WMD - weighted mean difference
- EEG - Electroencephalography
- Z-drugs - nonbenzodiazepine hypnotics

## Declarations

### Ethics approval and consent to participate

Not applicable. This study is a systematic review of previously published literature and does not involve direct contact with human or animal subjects. Therefore, ethical approval and patient consent were not needed.

### Consent for publication

Not Applicable

### Availability of data and materials

All meta-analytic data, the master data extraction spreadsheet, the completed risk of bias tools, and the analysis scripts (R) are publicly available with this manuscript at OpenICPSR https://www.openicpsr.org/openicpsr/project/247009/version/V1/view.

## Author contributions

Muhammad Awais Paracha (MAP), Sania Abdul Jalil Khan (SAJK), Rida Zarkaish (RZ), Faryal Fazal (FF), Muhammad Danish Khan (MDK), and Mahnoor Ahmad (MA) conceived this study. MAP and SAJ conducted the initial searches and performed the literature review. MAP, SAJK and FF contributed to protocol writing and registration at PROSPERO. All the authors contributed equally to screening the article via Rayyan without enabling the AI-assisted module. All the authors also contributed equally to the data extraction and risk of bias assessment. The MAP performed the meta-analysis using and oversaw the overall teamwork and offered regular feedback. MAP led the drafting and editing of the article. MAP, SAJK, RZ, FF, MDK, and MA drafted particular sections of the article. All the authors were involved in critically revising the article for important intellectual content. All authors approved the final version of the article. MAP is the guarantor of this work. The corresponding author attests that all listed authors meet authorship criteria and that no others meeting the criteria have been omitted.

## Supporting information

Search string

## Data Availability

All meta-analytic data, the master data extraction spreadsheet, the completed risk of bias tools, and the analysis scripts (R) are publicly available with this manuscript at OpenICPSR

https://www.openicpsr.org/openicpsr/project/247009/version/V1/view

## Acknowledgements

We acknowledge the Systematic Review Network and Dr. Yasith Mathangasinghe (Monash University) for their mentorship.

## Supplementary materials

**Appendix I: Search strategy**

**Appendix II: Tables**

Table 1: Study characteristics

Table 2: Study characteristics of the non-RCTs

Table 3: Results of continuous outcomes of individual RCTs

Table 4: Results of the continuous outcomes of individual non-RCTs

Table 5: GRADE summary of findings

**Appendix III: Figures**

## References

1. Benjafield AV, Sert Kuniyoshi FH, Malhotra A, Martin JL, Morin CM, Maurer LF, et al. Estimation of the global prevalence and burden of insomnia: a systematic literature review-based analysis. Sleep medicine reviews. 2025;82:102121.

2. Chokroverty S. Overview of sleep & sleep disorders. Indian Journal of Medical Research. 2010;131(2):126–40.

3. Brautsch LA, Lund L, Andersen MM, Jennum PJ, Folker AP, Andersen S. Digital media use and sleep in late adolescence and young adulthood: A systematic review. Sleep medicine reviews. 2023;68:101742.

4. Jaffer U, Sharminaz S, Zulkafli N, Omar M, Che Mohd Nassir CMN, Ahmed M, et al. Screen Time And Sleep Quality: A Narrative Review Of Digital Device Usage And Its Impact On Well-Being. International Journal of Education Psychology and Counselling. 2024;9:1068–79.

5. Arshad D, Joyia UM, Fatima S, Khalid N, Rishi AI, Rahim NUA, et al. The adverse impact of excessive smartphone screen-time on sleep quality among young adults: A prospective cohort. Sleep Science . 2021;14(4):337–41.

6. Zhong C, Masters M, Donzella SM, Diver WR, Patel AV. Electronic Screen Use and Sleep Duration and Timing in Adults. JAMA Network Open. 2025;8(3):e252493-e.

7. Cyr M, Artenie DZ, Al Bikaii A, Borsook D, Olson JA. The effect of evening light on circadian-related outcomes: A systematic review. Sleep medicine reviews. 2022 Aug 1;64:101660.

8. Gooley JJ, Chamberlain K, Smith KA, Khalsa SB, Rajaratnam SM, Van Reen E, Zeitzer JM, Czeisler CA, Lockley SW. Exposure to room light before bedtime suppresses melatonin onset and shortens melatonin duration in humans. J Clin Endocrinol Metab. 2011 Mar;96(3):E463–72. doi: 10.1210/jc.2010-2098. Epub 2010 Dec 30. PMID:21193540; PMCID: PMC3047226.

9. Green, A., Cohen-Zion, M., Haim, A., & Dagan, Y. (2017). Evening light exposure to computer screens disrupts human sleep, biological rhythms, and attention abilities. Chronobiology international, 34(7), 855–865. 10.1080/07420528.2017.1324878

10. Nakshine V S, Thute P, Khatib M, et al. (October 08, 2022) Increased Screen Time as a Cause of Declining Physical, Psychological Health, and Sleep Patterns: A Literary Review. Cureus 14(10): e30051. doi:10.7759/cureus.30051

11. Edinger JD, Means MK. Cognitive–behavioral therapy for primary insomnia. Clinical psychology review. 2005;25(5):539–58.

12. Holbrook AM, Crowther R, Lotter A, Cheng C, King D. Meta-analysis of benzodiazepine use in the treatment of insomnia. Cmaj. 2000;162(2):225–33.

13. Harbourt K, Nevo ON, Zhang R, Chan V, Croteau D. Association of eszopiclone, zaleplon, or zolpidem with complex sleep behaviors resulting in serious injuries, including death. Pharmacoepidemiology and drug safety. 2020;29(6):684–91.

14. U.S. Food and Drug Administration. FDA adds Boxed Warning for risk of serious injuries caused by sleepwalking with certain prescription insomnia medicines [Internet]. Silver Spring (MD): U.S. Food and Drug Administration; 2019 Apr 30 [cited 2026 Jan 7]. Available from: https://www.fda.gov/drugs/fda-drug-safety-podcasts/fda-adds-boxed-warning-risk-serious-injuries-caused-sleepwalking-certain-prescription-insomnia U.S. Food and Drug Administration+1

15. Salahub C, Wu PE, Burry LD, Soong C, Sheehan KA, MacMillan TE, et al. Melatonin for insomnia in medical inpatients: a narrative review. Journal of Clinical Medicine. 2022;12(1):256.

16. Sateia MJ, Buysse DJ, Krystal AD, Neubauer DN, Heald JL. Clinical practice guideline for the pharmacologic treatment of chronic insomnia in adults: an American Academy of Sleep Medicine clinical practice guideline. Journal of clinical sleep medicine. 2017;13(2):307–49.

17. Saleh AY, Saputra DAY, Valentina R, Susanto TD. How Extensively is Herbal Medicine Used as a Therapy for Insomnia? A Bibliometric Study. Pharmacogn J. 2025;17(3): 342–364

18. Leach, M. J., & Page, A. T. (2015). Herbal medicine for insomnia: A systematic review and meta-analysis. Sleep medicine reviews, 24, 1–12. 10.1016/j.smrv.2014.12.003

19. Patwardhan B, Warude D, Pushpangadan P, Bhatt N. Ayurveda and traditional Chinese medicine: a comparative overview. Evid Based Complement Alternat Med. 2005 Dec;2(4):465–73. doi: 10.1093/ecam/neh140. Epub 2005 Oct 27. PMID: 16322803; PMCID: PMC1297513.

20. Yeom JW, Cho CH. Herbal and Natural Supplements for Improving Sleep: A Literature Review. Psychiatry Investig. 2024 Aug;21(8):810–821. doi: 10.30773/pi.2024.0121. Epub 2024 Aug 2. PMID: 39086164; PMCID: PMC11321869.

21. Bent S. Herbal medicine in the United States: review of efficacy, safety, and regulation: grand rounds at University of California, San Francisco Medical Center. Journal of general internal medicine. 2008;23(6):854–9.

22. Calapai, G. (2008). European Legislation on Herbal Medicines: A Look into the Future. Drug Safety, 31(5), 428–431.

23. Bent S, Padula A, Moore D, Patterson M, Mehling W. Valerian for sleep: a systematic review and meta-analysis. The American journal of medicine. 2006;119(12):1005–12.

24. Kazemi, A., Shojaei-Zarghani, S., Eskandarzadeh, P., & Hashempur, M. H. (2024). Effects of chamomile (Matricaria chamomilla L.) on sleep: A systematic review and meta-analysis of clinical trials. Complementary therapies in medicine, 84, 103071. 10.1016/j.ctim.2024.103071

25. Kim GH, Kim Y, Yoon S, Kim SJ, Yi SS. Sleep-inducing effect of Passiflora incarnata L. extract by single and repeated oral administration in rodent animals. Food science & nutrition. 2020 Jan;8(1):557–66.

26. Oliveira MVB, Garguerra JA, Lamas CB, Laurindo LF, Rodrigues VD, Sloan KP, Sloan LA, Chagas EFB, Guiguer EL, Detregiachi CRP, et al. Unravelling the Effects of Melissa officinalis L. on Cognition and Sleep Quality: A Narrative Review. International Journal of Molecular Sciences. 2025; 26(21):10566. 10.3390/ijms262110566.

27. Guadagna S, Barattini DF, Rosu S, Ferini-Strambi L. Plant Extracts for Sleep Disturbances: A Systematic Review. Evid Based Complement Alternat Med. 2020 Apr 21;2020:3792390. doi: 10.1155/2020/3792390. PMID: 32382286; PMCID: PMC7191368.

28. Safari AA, Shirzad M, Mehraban MS, Kashani LM, Shams-Baghbanan H, Pourrostami K, Ahmadian-Attari MM. Sweet violet syrup reduces restlessness and improves appetite in children with fever: A double-blind randomized controlled trial. Advances in Integrative Medicine. 2025 Mar 7.

29. He S, Wang C, Shi W, Zhou C, Liu C, Xu Y, Gui Y, Jiang X, Fan G. Assessment of the Properties and Mechanism of Three Lotus (*Nelumbo nucifera* Gaertn.) Parts for Sleep Improvement. Chemistry & Biodiversity. 2025 Jun;22(6):e202402788

30. Becker A, Yamada Y, Sato F. California poppy (Eschscholzia californica), the Papaveraceae golden girl model organism for evodevo and specialized metabolism. Frontiers in plant science. 2023;14:1084358.

31. Sarris J, McIntyre E, Camfield DA. Plant-based medicines for anxiety disorders, part 1: a review of preclinical studies. CNS drugs. 2013;27(3):207–19.

32. Shevtsov V, Zholus B, Shervarly V, Vol’Skij V, Korovin Y, Khristich M, et al. A randomized trial of two different doses of a SHR-5 *Rhodiola rosea* extract versus placebo and control of capacity for mental work. Phytomedicine. 2003;10(2-3):95–105.

33. Sarris J, Panossian A, Schweitzer I, Stough C, Scholey A. Herbal medicine for depression, anxiety and insomnia: a review of psychopharmacology and clinical evidence. European neuropsychopharmacology. 2011;21(12):841–60.

34. Ernst E. Harmless herbs? A review of the recent literature. The American journal of medicine. 1998;104(2):170–8.

35. Poursaleh Z, Khodadoost M, Vahedi E, Attari M, Jafari M, Poursaleh E. A review of available herbal medicine options for the treatment of chronic insomnia. Pakistan Journal of Medical and Health Sciences. 2021;15:1589–98.

36. Li Y, Pham V, Bui M, Song L, Wu C, Walia A, Uchio E, Smith-Liu F, Zi X. *Rhodiola rosea* L.: an herb with anti-stress, anti-aging, and immunostimulating properties for cancer chemoprevention. Curr Pharmacol Rep. 2017 Dec;3(6):384–395. doi: 10.1007/s40495-017-0106-1. Epub 2017 Sep 14. PMID: 30393593; PMCID: PMC6208354.

37. Fedurco M, Gregorová J, Šebrlová K, Kantorová J, Peš O, Baur R, Sigel E, Táborská E. Modulatory Effects of Eschscholzia californica Alkaloids on Recombinant GABAA Receptors. Biochem Res Int. 2015;2015:617620. doi: 10.1155/2015/617620. Epub 2015 Oct 5. PMID: 26509084; PMCID: PMC4609799.

38. Page MJ, McKenzie JE, Bossuyt PM, Boutron I, Hoffmann TC, Mulrow CD, et al. The PRISMA 2020 statement: an updated guideline for reporting systematic reviews. BMJ 2021;372:n71. doi: 10.1136/bmj.n71. For more information, visit: https://www.prisma-statement.org/

39. Ouzzani, M., Hammady, H., Fedorowicz, Z. et al. Rayyan a web and mobile app for systematic reviews. Syst Rev 5, 210 (2016). 10.1186/s13643-016-0384-4

40. Cochrane Effective Practice and Organization of Care (EPOC). Data collection form. EPOC Resources for review authors, 2017. epoc.cochrane.org/resources/epoc-specific-resources-review-authors [accessed July 15, 2025

41. Sterne JAC, Savović J, Page MJ, Elbers RG, Blencowe NS, Boutron I, et al. RoB 2: a revised tool for assessing risk of bias in randomized trials. BMJ. 2019;366:l4898. doi:10.1136/bmj.l4898

42. Sterne JAC, Hernán MA, Reeves BC, Savović J, Berkman ND, Viswanathan M, Henry D, Altman DG, Ansari MT, Boutron I, Carpenter JR, Chan AW, Churchill R, Deeks JJ, Hróbjartsson A, Kirkham J, Jüni P, Loke YK, Pigott TD, Ramsay CR, Regidor D, Rothstein HR, Sandhu L, Santaguida PL, Schünemann HJ, Shea B, Shrier I, Tugwell P, Turner L, Valentine JC, Waddington H, Waters E, Wells GA, Whiting PF, Higgins JPT. ROBINS-I: a tool for assessing risk of bias in non-randomized studies of interventions. BMJ 2016; 355; i4919; doi: 10.1136/bmj.i4919.

43. Huang, XT, Automatic tool for Risk of Bias In Nonrandomized Studies of Interventions (ROBINS-I), (2025) National Drug and Alcohol Research Centre (NDARC), University of New South Wales, Australia. Available from: https://www.unsw.edu.au/research/ndarc/resources/risk_of_bias_tool

44. McGuinness, LA, Higgins, JPT. Risk-of-bias VISualization (robvis): An R package and Shiny web app for visualizing risk-of-bias assessments. Res Syn Meth. 2020; 1–7. 10.1002/jrsm.1411

45. Schünemann, H. J., Higgins, J. P. T., Vist, G. E., Glasziou, P., Akl, E. A., Skoetz, N., & Guyatt, G. H. (2019). Chapter 15: Interpreting results and drawing conclusions. In J. P. T. Higgins, J. Thomas, J. Chandler, M. Cumpston, T. Li, M. J. Page, & V. A. Welch (Eds.), Cochrane Handbook for Systematic Reviews of Interventions (2nd ed.). Wiley. https://training.cochrane.org/handbook/current/chapter-15

46. Campbell M, McKenzie J E, Sowden A, Katikireddi S V, Brennan S E, Ellis S et al. Synthesis without meta-analysis (SWiM) in systematic reviews: reporting guideline BMJ 2020; 368 :l6890 doi:10.1136/bmj.l689

47. Higgins JPT, Thomas J, Chandler J, Cumpston M, Li T, Page MJ, et al., Cochrane Handbook for Systematic Reviews of Interventions version 6.5 (updated August 2024) Chapter 14: Completing ‘Summary of findings’ tables and grading the certainty of the evidence. Cochrane, 2024. Available from www.cochrane.org/handbook

48. Balshem, H., Helfand, M., Schünemann, H. J., Oxman, A. D., Kunz, R., Brozek, J., et al., 2011. GRADE guidelines: 3. Rating the quality of evidence. Journal of clinical epidemiology, 64(4), 401–406. 10.1016/j.jclinepi.2010.07.015

49. Evidence Prime. (2015). GRADEpro Guideline Development Tool [Software]. McMaster University. https://www.gradepro.org

50. Abdullahzadeh, M., Matourypour, P., & Naji, S. A. (2017). Investigation effect of oral chamomilla on sleep quality in elderly people in Isfahan: A randomized control trial. Journal of education and health promotion, 6, 53. Available from: 10.4103/jehp.jehp_109_15

51. Zick, S. M., Wright, B. D., Sen, A., & Arnedt, J. T. (2011). Preliminary examination of the efficacy and safety of a standardized chamomile extract for chronic primary insomnia: a randomized placebo-controlled pilot study. BMC complementary and alternative medicine, 11, 78. Available from: 10.1186/1472-6882-11-78

52. Abdullahzadeh M, Naji SA. (2014).The effect of matricaria chamomilla on sleep quality of elderly people admitted to nursing homes. Iran Journal of Nursing Vol. 27, no. 89. pp. 69–79 Available from: 10.29252/ijn.27.89.69

53. Abbasinia H, Alizadeh Z, Vakilian K, Jafari Z, Ghorbani M, Shariat M. Effect of Chamomile extract on sleep disorder in menopausal women. *Iranian Journal of Obstetrics*, Gynecology and Infertility. 2016 Jan; 19(20):1–7. Available from: https://www.doi.org/10.22038/ijogi.2016.7631

54. Jarukitsakul S, Keawjeen T, Inthanuchit K, Chatawatee B. (2025). A Comparative Study on the Effectiveness of Pepper Leaves Tea and Chamomile Tea for Insomnia Disorder: A Single-Blind Randomized Clinical Trial. Tropical Journal of Natural Product Reserach May;9(5):1977. Available from: https://www.doi.org/10.26538/tjnpr/v9i5.14

55. Alvarado-Garcia PAA, Soto-Vásquez MR, Rodrigo-Villanueva EM, Gavidia-Valencia JG, Rodríguez-Guzma NM, Rengifo-Penadillos RA, Campos-Florián JV, Rodríguezde Guzmán YE. (2024) Chamomile (Matricaria chamomilla L.). Essential Oil and its Potential Against Stress, Anxiety, and Sleep Quality. Pharmacognosy Journal, 16(1): 101–107 Available from: http://www.phcogj.com/v16/i1

56. Chang, S. M., & Chen, C. H. (2016). Effects of an intervention with drinking chamomile tea on sleep quality and depression in sleep disturbed postnatal women: a randomized controlled trial. Journal of advanced nursing, 72(2), 306–315. Available from: 10.1111/jan.12836

57. Adib-Hajbaghery, M., & Mousavi, S. N. (2017). The effects of chamomile extract on sleep quality among elderly people: A clinical trial. Complementary therapies in medicine, 35, 109–114. Available from: 10.1016/j.ctim.2017.09.010

58. Deepa, Y., Vijay, A., Nivethitha, L., Nandhakumar, G., Sathiya, S., & Mooventhan, A. (2025). Effects of chamomile oil inhalation on sleep quality in young adults with insomnia: A randomized controlled trial. International journal of psychiatry in medicine, 60(5), 533–542. Available from: 10.1177/00912174241301279

59. Renosih NE, Nisa MK, Dawis AM 2025. The impact of a bergamot and chamomile aromatherapy blend on sleep quality in elderly individuals with insomnia. International Journal of Islamic and Complementary Medicine Jan;6(1):26–30. Available from: https://www.doi.org/10.55116/IJICM.V6I1.103

60. Harit MK, Mundhe N, Tamoli S Sr, Pawar V, Bhapkar V, Kolhe G, Mahadik S, Kulkarni A, Agarwal A. 2024 Randomized, Double-Blind, Placebo-Controlled, Clinical Study of Passiflora incarnata in Participants With Stress and Sleep Problems. Cureus. Mar 20;16(3):e56530. Available from: https://www.doi.org/10.7759/cureus.56530

61. Lee, J., Jung, H. Y., Lee, S. I., Choi, J. H., & Kim, S. G. (2020). Effects of Passiflora incarnata Linnaeus on polysomnographic sleep parameters in subjects with insomnia disorder: a double-blind randomized placebo-controlled study. International clinical psychopharmacology, 35(1), 29–35. Available from: 10.1097/YIC.0000000000000291

62. Ngan, A., & Conduit, R. (2011). A double-blind, placebo-controlled investigation of the effects of Passiflora incarnata (passionflower) herbal tea on subjective sleep quality. Phytotherapy research : PTR, 25(8), 1153–1159. Available from: 10.1002/ptr.3400

63. Marcos E, Iglesias I, Vazquez-Velasco M, Benedi J. Community pharmacy-based interventions with Valeriana officinalis or Passiflora incarnata together with sleep hygiene education improve climacteric symptoms and sleep problems in menopause. JONNPR. 2020;5(12):1538–57 Available from: 10.19230/jonnpr.3983

64. Taavoni, S., Ekbatani, N.N., & Haghani, H. (2013). The Effect of lemon Balm on sleep disorder in menopausal women 60-50 years old. Complementary Medicine Journal of Faculty of Nursing and Midwifery, 2, 344–354. Available from: https://cmja.arakmu.ac.ir/browse.php?a_id=98&sid=1&slc_lang=en

65. Pierro, F., Sisti, D., Rocchi, M., Belli, A., Bertuccioli, A., Cazzaniga, M., Palazzi, C. M., Tanda, M. L., & Zerbinati, N. (2024). Effects of *Melissa officinalis* Phytosome on Sleep Quality: Results of a Prospective, Double-Blind, Placebo-Controlled, and Cross-Over Study. Nutrients, 16(23), 4199. Available from: 10.3390/nu16234199

66. Ranjbar, M., Salehi, A., Rezaeizadeh, H., Zarshenas, M. M., Sadeghniiat-Haghighi, K., Mirabzadeh, M., & Firoozabadi, A. (2018) a. Efficacy of a Combination of Melissa officinalis L. and Nepeta Menthoides Boiss. & Buhse on Insomnia: A Triple-Blind, Randomized Placebo-Controlled Clinical Trial. Journal of alternative and complementary medicine (New York, N.Y.), 10.1089/acm.2017.0153. Advance online publication. Available from: https://doi.org/10.1089/acm.2017.0153

67. Ranjbar, M., Firoozabadi, A., Salehi, A., Ghorbanifar, Z., Zarshenas, M. M., Sadeghniiat-Haghighi, K., & Rezaeizadeh, H. (2018)b. Effects of Herbal combination (Melissa officinalis L. and Nepeta menthoides Boiss. & Buhse) on insomnia severity, anxiety and depression in insomniacs: Randomized placebo controlled trial. Integrative medicine research, 7(4), 328–332. Available from: 10.1016/j.imr.2018.08.001

68. Feyzabadi, Z., Rezaeitalab, F., Badiee, S., Taghipour, A., Moharari, F., Soltanifar, A., & Ahmadpour, M. R. (2018). Efficacy of Violet oil, a traditional Iranian formula, in patients with chronic insomnia: A randomized, double-blind, placebo-controlled study. Journal of ethnopharmacology, 214, 22–28. Available from: 10.1016/j.jep.2017.11.036

69. Taherzadeh, Z., Khaluyan, H., Iranshahy, M., Rezaeitalab, F., Eshaghi Ghalibaf, M. H., & Javadi, B. (2020). Evaluation of sedative effects of an intranasal dosage form containing saffron, lettuce seeds and sweet violet in primary chronic insomnia: A randomized, double-dummy, double-blind placebo controlled clinical trial. Journal of ethnopharmacology, 262, 113116. Available from: 10.1016/j.jep.2020.113116

70. Maroo, N., Hazra, A., & Das, T. (2013). Efficacy and safety of a polyherbal sedative-hypnotic formulation NSF-3 in primary insomnia in comparison to zolpidem: a randomized controlled trial. Indian journal of pharmacology, 45(1), 34–39. Available from: 10.4103/0253-7613.106432

71. Gutiérrez-Romero, S. A., Torres-Narváez, E. S., Zamora-Gómez, A. C., Castillo-Castillo, S., Latorre-Velásquez, A. L., Betancourt-Villamizar, C., & Mendivil, C. O. (2024). Effect of a nutraceutical combination on sleep quality among people with impaired sleep: a randomized, placebo-controlled trial. Scientific reports, 14(1), 8062. Available from: 10.1038/s41598-024-58661-z

72. Bongartz, U., Tan, B. K., Seibt, S., Bothe, G., Uebelhack, R., Chong, P. W., & Wszelaki, N. (2019). Sleep Promoting Effects of IQP-AO-101: A Double-Blind, Randomized, Placebo-Controlled Exploratory Trial. Evidence-based complementary and alternative medicine : eCAM, 2019, 9178218. 10.1155/2019/9178218

73. Abdellah, S. A., Berlin, A., Blondeau, C., Guinobert, I., Guilbot, A., Beck, M., & Duforez, F. (2019). A combination of *Eschscholtzia californica* Cham. and *Valeriana officinalis* L. extracts for adjustment insomnia: A prospective observational study. Journal of traditional and complementary medicine, 10(2), 116–123. Available from: 10.1016/j.jtcme.2019.02.003

74. Catherin, M.DE, Prabavathy S, Renuka K (2021). Effect of chamomile oil on stress, anxiety and quality of sleep among elderly in selected old age home at Puducherry. International Journal of Current Research. Dec;13(12):19868–72. Available from: https://www.doi.org/10.24941/ijcr.42693.12.2021

75. Feyzabadi Z, Jafari F, Kamali SH, Ashayeri H, Badiee Aval S, Esfahani MM, Sadeghpour O. Efficacy of Viola odorata in Treatment of Chronic Insomnia. Iran Red Crescent Medical Journal. 2014 Dec 14;16(12):e17511. Available from: 10.5812/ircmj.17511

76. Cases J, Ibarra A, Feuillère N, Roller M, Sukkar SG. Pilot trial of Melissa officinalis L. leaf extract in the treatment of volunteers suffering from mild-to-moderate anxiety disorders and sleep disturbances. The Mediterranean Journal of Nutrition and Metabolism. 2011 Dec;4(3):211–218. Available from: 10.1007/s12349-010-0045-4

77. Kim, Y., Lee, W. K., Jeong, H., Choi, H. J., Lee, M. K., & Cho, S. H. (2024). Mixture of *Rhodiola rosea* and *Nelumbo nucifera* Extracts Ameliorates Sleep Quality of Adults with Sleep Disturbance. Nutrients, 16(12), 1867. Available from: 10.3390/nu16121867

78. Simone, M., De Feo, R., Choucha, A., Ciaglia, E., & Fezeu, F. (2023). Enhancing Sleep Quality: Assessing the Efficacy of a Fixed Combination of Linden, Hawthorn, Vitamin B1, and Melatonin. Medical sciences (Basel, Switzerland), 12(1), 2. Available from: 10.3390/medsci12010002

79. Kimtata V, Ahmed HA, Sushma B. 2023. A prospective, open-label, nonrandomized study to assess the efficacy and safety of zero tension tablets in patients with stress, anxiety and insomnia disorders. International Journal of Research in Ayurveda and Pharmacy. Apr;14(2):80–5. Available from: https://www.doi.org/10.7897/2277-4343.140246

80. Lemoine, P., Bablon, J. C., & Da Silva, C. (2019). A combination of melatonin, vitamin B6 and medicinal plants in the treatment of mild-to-moderate insomnia: A prospective pilot study. Complementary therapies in medicine, 45, 104–108. Available from: 10.1016/j.ctim.2019.05.024

81. Jaroenngarmsamer P, Pajongsaleepanya K, Rojanabenjakun P, Ounprasertsuk J, Srijaruneruang S, Thongkham K, Kaewmanee T. 2021 .Efficiency of chamomile essential oils on sleeping quality of first-year university students. Indian journal of Forensic Medicine & Toxicology Mar;15(2):3724–31. Available from: https://www.doi.org/10.37506/ijfmt.v15i2.14954

82. Khalid, Z., Noreen, S., Aja, P. M., Rehman, A., & Sadiqa, A. (2025). Comparative effect of Chamomile flower (*Matricaria chamomilla* L.) and Passion flower (*Passiflora incarnata* L.) powder tea in patients suffering from primary insomnia. CyTA - Journal of Food, 23(1). 10.1080/19476337.2025.2504527

83. Kazemi, A., Shojaei-Zarghani, S., Eskandarzadeh, P., & Hashempur, M. H. (2024). Effects of chamomile (Matricaria chamomilla L.) on sleep: A systematic review and meta-analysis of clinical trials. Complementary therapies in medicine, 84, 103071. 10.1016/j.ctim.2024.103071

84. Srivastava, J.K., Shankar, E., & Gupta, S. (2010). Chamomile: A herbal medicine of the past with a bright future (Review). Molecular Medicine Reports, 3, 895–901 10.3892/mmr.2010.377

85. Yurcheshen, M., Seehuus, M., & Pigeon, W. (2015). Updates on Nutraceutical Sleep Therapeutics and Investigational Research. Evidence-based complementary and alternative medicine : eCAM, 2015, 105256. 10.1155/2015/105256

86. Yeom, J. W., & Cho, C. H. (2024). Herbal and Natural Supplements for Improving Sleep: A Literature Review. Psychiatry investigation, 21(8), 810–821. 10.30773/pi.2024.0121

87. National Center for Complementary and Integrative Health. (NCCIH) Chamomile [Internet]. Bethesda (MD): National Institutes of Health; 2024 Nov [cited 2026 Feb 1]. Available from: https://www.nccih.nih.gov/health/chamomile

88. Shinomiya, K., Inoue, T., Utsu, Y., Tokunaga, S., Masuoka, T., Ohmori, A., & Kamei, C. (2005). Hypnotic activities of chamomile and passiflora extracts in sleep-disturbed rats. Biological & pharmaceutical bulletin, 28(5), 808–810. 10.1248/bpb.28.808

89. Miraj S, Alesaeidi S. A systematic review study of therapeutic effects of Matricaria recuitta chamomile (chamomile). Electron Physician. 2016 Sep 20;8(9):3024–3031 10.19082/3024

90. Winkler, A., Auer, C., Doering, B. K., & Rief, W. (2014). Drug treatment of primary insomnia: a meta-analysis of polysomnographic randomized controlled trials. CNS drugs, 28(9), 799–816. 10.1007/s40263-014-0198-7

91. Oliveira, M. V. B., Garguerra, J. A., Lamas, C. B., Laurindo, L. F., Rodrigues, V. D., Sloan, K. P., Sloan, L. A., Chagas, E. F. B., Guiguer, E. L., Detregiachi, C. R. P. Miglino, M. A., Pereira, E. S. B. M., Valenti, V. E., Silva, L. R., & Barbalho, S. M. (2025). Unravelling the Effects of Melissa officinalis L. on Cognition and Sleep Quality: A Narrative Review. International journal of molecular sciences, 26(21), 10566. 10.3390/ijms262110566

92. Soltanpour A, Alijaniha F, Naseri M, Kazemnejad A, Heidari MR. Effects of Melissa officinalis on anxiety and sleep quality in patients undergoing coronary artery bypass surgery: A double-blind randomized placebo controlled trial. European Journal of Integrative Medicine. 2019 Jun 1;28:27–32. 10.1016/j.eujim.2019.01.010

93. Wang C-C, Hsieh P-W, Kuo J-R, Wang S-J. Rosmarinic Acid, a Bioactive Phenolic Compound, Inhibits Glutamate Release from Rat Cerebrocortical Synaptosomes through GABAA Receptor Activation. Biomolecules. 2021; 11(7):1029. 10.3390/biom11071029

94. Guerrero, F. A., & Medina, G. M. (2017). Effect of a medicinal plant (Passiflora incarnata L) on sleep. Sleep science (Sao Paulo, Brazil), 10(3), 96–100. 10.5935/1984-0063.20170018

95. Kim, G. H., Kim, Y., Yoon, S., Kim, S. J., & Yi, S. S. (2019). Sleep-inducing effect of *Passiflora incarnata L.* extract by single and repeated oral administration in rodent animals. Food science & nutrition, 8(1), 557–566. 10.1002/fsn3.1341

96. Lee, J., Jung, H. Y., Lee, S. I., Choi, J. H., & Kim, S. G. (2020). Effects of Passiflora incarnata Linnaeus on polysomnographic sleep parameters in subjects with insomnia disorder: a double-blind randomized placebo-controlled study. International clinical psychopharmacology, 35(1), 29–35. 10.1097/YIC.0000000000000291

97. La Tempa, A.; Ferraiuolo, G.; Pranzetti, B.; Pruccoli, J.; Parmeggiani, A. Passiflora Incarnata L. Herba in the Treatment of Anxiety Symptoms and Insomnia in Children and Adolescents with Feeding and Eating Disorders. Adolescents 2025, 5, 24. 10.3390/adolescents5020024

98. Huang, S., Huang, Q., Zhou, Z., Zhang, J., Zhan, Y., & Liang, Z. (2022). The Efficacy of V. odorata Extract in the Treatment of Insomnia: A Systematic Review and Meta-Analysis. Frontiers in neurology, 13, 730311. 10.3389/fneur.2022.730311

99. Ahn, Y., Kim, S., Park, C., Kim, J. E., Suh, H. J., & Jo, K. (2022). Sleep-promoting activity of lotus (*Nelumbo nucifera*) rhizome water extract via GABAA receptors. Pharmaceutical biology, 60(1), 1341–1348. 10.1080/13880209.2022.2096076

100. Yu, H. L., Zhang, P. P., Zhang, C., Zhang, X., Li, Z. Z., Li, W. Q., & Fu, A. S. (2019). Lin chuang er bi yan hou tou jing wai ke za zhi = Journal of clinical otorhinolaryngology head and neck surgery, 33(10), 954–957. 10.13201/j.issn.1001-1781.2019.10.013

101. Ishaque, S., Shamseer, L., Bukutu, C. et al. *Rhodiola rosea* for physical and mental fatigue: a systematic review. BMC Complement Altern Med 12, 70 (2012). 10.1186/1472-6882-12-70

102. Panossian, A., Wikman, G., & Sarris, J. (2010). Rosenroot (*Rhodiola rosea*): traditional use, chemical composition, pharmacology and clinical efficacy. Phytomedicine : international journal of phytotherapy and phytopharmacology, 17(7), 481–493. 10.1016/j.phymed.2010.02.002

103. Anghelescu, I. G., Edwards, D., Seifritz, E., & Kasper, S. (2018). Stress management and the role of *Rhodiola rosea*: a review. International journal of psychiatry in clinical practice, 22(4), 242–252. 10.1080/13651501.2017.1417442

104. Fedurco, M., Gregorová, J., Šebrlová, K., Kantorová, J., Peš, O., Baur, R., Sigel, E., & Táborská, E. (2015). Modulatory Effects of Eschscholzia californica Alkaloids on Recombinant GABAA Receptors. Biochemistry research international, 2015, 617620. 10.1155/2015/617620

105. Arora A. A study to compare the efficacy of Eschscholzia californica MT and Passiflora incarnata MT in insomnia. International Journal of Homoeopathic Sciences. 2019;3(03):18–22. 10.33545/26164485.2019.v3.i3a.89

106. Abbasi, M., Gohari, S., Ahangar, H., Bahrami, M., Kamalinejad, M., Reshadmanesh, T., & Nazari, S. S. (2021). Efficacy of Hawthorn fruit extract on blood pressure and quality of sleep in patients with hypertension along with sleep disorders: A randomized double-blind controlled trial. Journal of Contemporary Medical Sciences, 7(4), 196–201. 10.22317/jcms.v7i4.1017

107. Deodhar M, Al Rihani SB, Arwood MJ, Darakjian L, Dow P, Turgeon J, Michaud V. Mechanisms of CYP450 Inhibition: Understanding Drug Drug Interactions Due to Mechanism-Based Inhibition in Clinical Practice. Pharmaceutics. 2020 Sep 4;12(9):846. 10.3390/pharmaceutics12090846

108. Buysse, D. J., Hall, M. L., Strollo, P. J., Kamarck, T. W., Owens, J., Lee, L., Reis, S. E., & Matthews, K. A. (2008). Relationships between the Pittsburgh Sleep Quality Index (PSQI), Epworth Sleepiness Scale (ESS), and clinical/polysomnographic measures in a community sample. Journal of clinical sleep medicine : JCSM : official publication of the American Academy of Sleep Medicine, 4(6), 563–571.

109. Sarris, J., Panossian, A., Schweitzer, I., Stough, C., & Scholey, A. (2011). Herbal medicine for depression, anxiety and insomnia: a review of psychopharmacology and clinical evidence. European neuropsychopharmacology : the journal of the European College of Neuropsychopharmacology, 21(12), 841–860. 10.1016/j.euroneuro.2011.04.002

110. Zhao, F. Y., Xu, P., Kennedy, G. A., Conduit, R., Zhang, W. J., Wang, Y. M., Fu, Q. Q., & Zheng, Z. (2023). Identifying complementary and alternative medicine recommendations for insomnia treatment and care: a systematic review and critical assessment of comprehensive clinical practice guidelines. Frontiers in public health, 11, 1157419. 10.3389/fpubh.2023.1157419

111. Ng, J. Y., & Parakh, N. D. (2021). A systematic review and quality assessment of complementary and alternative medicine recommendations in insomnia clinical practice guidelines. BMC complementary medicine and therapies, 21(1), 54. 10.1186/s12906-021-03223-3

112. Ekor M. The growing use of herbal medicines: issues relating to adverse reactions and challenges in monitoring safety. Front Pharmacol. 2014 Jan 10;4:177 10.3389/fphar.2013.00177

113. Hess D. R. (2023). Observational Studies. Respiratory care, 68(11), 1585–1597. 10.4187/respcare.11170

114. OCEBM Levels of Evidence Working Group (2011) The Oxford Levels of Evidence 2. Oxford: Oxford Centre for Evidence-Based Medicine. Available at: https://www.cebm.ox.ac.uk/resources/levels-of-evidence/ocebm-levels-of-evidence [Accessed 1 February 2026].

115. Spanakis M, Bakaros E, Papadopoulou S N, Fournaraki A, Symvoulakis EK. Pharmacoepidemiological Data on Drug-Herb Interactions: Serotonin Syndrome, Arrhythmias and the Emerging Role of Artificial Intelligence. Pharmacoepidemiology. 2025; 4(4):22. 10.3390/pharma4040022

116. Hassen, G., Belete, G., Carrera, K. G., Iriowen, R. O., Araya, H., Alemu, T., Solomon, N., Bam, D. S., Nicola, S. M., Araya, M. E., Debele, T., Zouetr, M., & Jain, N. (2022). Clinical Implications of Herbal Supplements in Conventional Medical Practice: A US Perspective. Cureus, 14(7), e26893. 10.7759/cureus.26893

117. Gagnier, J. J., Boon, H., Rochon, P., Moher, D., Barnes, J., Bombardier, C., & CONSORT Group (2006). Recommendations for reporting randomized controlled trials of herbal interventions: Explanation and elaboration. Journal of clinical epidemiology, 59(11), 1134–1149. 10.1016/j.jclinepi.2005.12.020.

118. Bruni, O., Ferini-Strambi, L., Giacomoni, E., & Pellegrino, P. (2021). Herbal Remedies and Their Possible Effect on the GABAergic System and Sleep. Nutrients, 13(2), 530. 10.3390/nu13020530

