## Supplementary material for "Effectiveness of Lesser Known Herbal Sedatives for Insomnia: A Systematic Review and Meta-Analysis": Table

**Appendix II: Tables**

**Table 1: Study Characteristics of RCTs**

| **Author (Year)** | **Country** | **No of Participants** | **Age Range** | **Primary clinical characteristics** | **Secondary clinical characteristics** | **Intervention 1/ Frequency** | **Intervention 2** | **Control** | **Primary Outcome** | **Secondary Outcome** | **Assessment time**  **T1-T7** | **Data** |
| --- | --- | --- | --- | --- | --- | --- | --- | --- | --- | --- | --- | --- |
| Abdullahzadeh et al., 2017 [50] | Iran | 77 | 60 years or older | Elderly people (age 60 and above) with sleep disorders | NR | Chamomile extract capsules/ 4 weeks | NA | Control | Pittsburgh Sleep Quality Index (PSQI) | NA | Baseline, 4 weeks after intervention | M, SD |
| Zick et al., 2011 [51] | USA | 34 | 18-65 | DSM-IV primary insomnia for ≥ 6 months | NR | Chamomile High Grade Extract (MediHerb, Warwick, Australia)/ 4 weeks (28 days) | NA | Placebo | Pittsburgh Sleep Quality Index (PSQI), Insomnia Severity Index (ISI), Total Sleep Time (hrs), Sleep Efficiency | NA | Baseline, 4 weeks | M, SD |
| Abdollahzadeh and Naji, 2014 [52] | Iran | 77 | 64 years and above | Elderly population | NR | Chamomille Extarct/ 8 weeks | NA | Placebo/ 8 weeks | Pittsburgh Sleep Quality Index (PSQI), | NR | Baseline, after 8 weeks | M, SD |
| Abbasinia et al, 2016 [53] | Iran | 110 | 51-61 | Menopausal women | Sleep disorders | Chamomile capsules/ 4 weeks | NA | Control | Pittsburgh Sleep Quality Index (PSQI), | NR | Baseline, after 4 weeks | M, SD |
| Jarukitsakul et al,2025 [54] | Thailand | 30 | 20-60 | Insomnia severity | Stress levels | Pepper Leaves Tea/ 3 weeks | NA | Chamomile Tea | ISI, PSQI, | Level of stress, satisfaction, safety assessment | Baseline, 7^th^ day, 14^th^ day, 21^st^ day | M, SD |
| Alvarado-Garcia, 2024 [55] | Peru | 110 | 18-45 years | Adults (male and female) experiencing stress, anxiety, and poor sleep quality | NA | Chamomile (Matricaria chamomilla L.) Essential Oil Aromatherapy/ 8 weeks | NA | Placebo | PSQI, SAS, | NA | Baseline, 8 weeks | M, SD |
| Chang and Chen, 2016 [56] | Taiwan | 80 | 24-43 | Postnatal women with poor sleep quality | Depression and Fatigue | Chamomile Tea Intervention/ 2 weeks | NA | Routine Postpartum Care | PSQI, EPDS, PFS | Infant night-care-related daytime dysfunction (PSQS subscale | Baseline, 2 weeks, 4 weeks | M, SD |
| Adib-Hajbaghery et al,2017 [57] | Iran | 40 | 60 years or more. | Elderly individuals | Some had chronic comorbidities (gastritis, arthritis, hypertension, Type II diabetes); hypnotic drug use noted in baseline (20% treatment, 13.3% control) | Chamomile extract capsules/ 4 weeks | NA | Placebo capsules | PSQI, | Components of PSQI | Baseline, 2 weeks, 4 weeks | M, SD |
| Deepa et al, 2024 [58] | India | 80 | 18-35 years | Insomnia and poor sleep quality | NR | Chamomile Oil Inhalation (Aromatherapy Group)/ 2 weeks | NA | Control | PSQI, ISI, | NA | Baseline, 2 weeks, | M, SD |
| Renosih et al,2025 [59] | Indonesia | NG | 60-75 years | PSQI | NR | a combination of bergamot aromatherapy and chamomile tea/ 4 weeks | NA | Control | PSQI | NA | Baseline, 4 weeks | M, SD |
| Harit et al., 2024 [60] | India | 65 | 18-60 | Insomnia (ISI 7-21) | Perceived Stress PSS score | SIVI (Passiflora incarnata extract)/ 4 weeks | NA | Placebo capsules (microcrystalline cellulose), taken at bedtime for 30 days. | Total sleep time. PSS | PSQI, GHQ-12, ISI, | Baseline, day 15, 30 day | M, SD |
| Lee et al, 2019 [61] | Republic of Korea | 80 | 18-59 | Adults aged 18-59 years diagnosed with insomnia disorder according to DSM-5 criteria. | Self-reported measures of depression (CES-D), anxiety (BAI), stress (GARS), and subjective sleep parameters (ISI, PSQI). | Passiflora incarnata (Passionflower) extract/ 2 weeks | NA | Placebo | TST, Sleep Efficiency (%),Sleep Latency, Wake After Sleep Onset (WASO, min) | ISI, PSQI, Center for Epidemiologic Studies Depression Scale (CES-D), BAI | Baseline, 2 weeks | M, SD |
| Ngan and Conduit, 2011 [62] | Australia | 41 | 18-35 | Healthy volunteers | NA | Passiflora incarnata (Passionflower) herbal tea/ 1 week | NA | Placebo Herbal Tea | Subjective Sleep Quality, Sleep Efficiency (%), Total sleep time | NA | Baseline, 1 week | M, SD |
| Marcos et al., 2020 [63] | Spain | 132 female | 45-64 | Menopausal women with sleep disturbance | Vasomotor and physical symptoms, insomnia severity, daytime sleepiness, quality of life satisfaction. | Passiflora incarnata (PI Group)/ 12 weeks | Valeriana officinalis (VO Group)/ 12 weeks | Sleep Hygiene Instructions Only (SHI Group)/ 12 weeks | PSQI, ISI, ESS, | Change in mental health–related quality of life, Treatment Satisfaction Questionnaire for Medication | Baseline, 4 weeks, 12 weeks | M, SD |
| Taavoni et. al, 2013 [64] | Iran | 100 females | 50-60 | Menopausal women | Sleep disorders | Lemon Balm/ 4 weeks | NA | Placebo | PSQI | NR | Baseline, after 4 weeks | M, SD |
| Pierro et al, 2024 [65] | Italy | 30 | 18-65 | perception of fatigue upon waking and unrefreshing sleep | Tolerability,compliance, physical activity, Anxiety level, side effects | Melissa officinalis Phytosome (MOP) supplementation/ 2 weeks | NA | Placebo | sleep quality and sleep architecture | SSQ, Anxiety level, physical activity level | Baseline, after 2 weeks first, 2 weeks second | M, SD |
| Ranjbar et al, 2018a [66] | Iran | 67 | 18 -60 | Insomnia Severity | Sleep quality and sleep time | Officinalis plus 400mg of N.menthoides/ 4 weeks | NA | Placebo | ISI, PSQI, | Sleep diary, Total Sleep time | Baseline, 2^nd^ week, 4^th^ week | M, SD |
| Ranjbar et al, 2018b [67] | Iran | NG | 18 -60 | ISI, BAI, BDI | NA | Herbal combination of Melissa officinalis (lemon balm) and Nepeta menthoides (Persian lavender/ 4 weeks | NA | Placebo | ISI, BDI, BAI, | NA | Baseline, 2 week, 4 week | M, SD |
| Feyzabadi et al., 2017 [68] | Iran | 60 | 18-60 | ISI | Sleep quality | Violet oil/ 4 weeks | Almond oil/ 4 weeks | Placebo | ISI, PSQI, | NA | Baseline, 30 days | M, SD |
| Taherzadeh et al., 2020 [69] | Iran | 66 | 18-40 | ISI | Sleep quality | Herbal oil/ 8 weeks | NA | Placebo (Sesame oil)/ 8 weeks | ISI | Sleep quality, sleep onset latency, Total sleep time | Baseline, 1 week. 4 week, 8 week | M, SD |
| Maroo et al, 2013 [70] | India | 78 | 20-80 | Sleep Problems | NA | NSF-3 (polyherbal sedative-hypnotic formulation)/ 2 weeks | NA | Zolpidem (Control Group) a benzodiazepine-like sedative-hypnotic drug/ 2 weeks | ISI, Total sleep time, Night time awakenings | ESS, Treatment-emergent adverse events (AEs) | Baseline, 1 week, 2 week | M, SD |
| Gutiérrez-Romero et al., 2024 [71] | Colombia | 64 | 18 and above | Poor sleep quality | NA | Nutraceutical combination (Green tea, Lemon balm, Valerian, and Saffron extract)/ 6 weeks | NA | Placebo Powder Mixture | PSQI, Sleep Latency, Sleep duration, Sleep Efficiency | Daytime dysfunction | Baseline, 6 weeks | M, SD |
| Bongartz et al., 2019 [72] | Germany | 50 | 23-64 | Poo sleep quality and insomnia symptoms | NA | IQP-AO-101 (Botanical Vitamin Complex)/ 6 weeks | NA | Placebo | Modified Athens Insomnia Scale (mAIS), Night Parameters, | NA | Baseline, 1 week, 4 week, 6 week | M, SD |

**Abbreviations:** PSQI, Pittsburg sleep quality Index; PSS, Perceived Stress Scale; ISI, Insomnia severity index; BAI, Beck Anxiety Inventory; BDI, Beck Depression Inventory; SAS, Self-rating Anxiety Scale; ESS, Epworth Sleepiness scale; EPDS, Edinburgh Postnatal depression scale; PFS, Parkinson Fatigue Scale; M, Mean; SD, Standard deviation; DSM, Diagnostic Statistical Manual of Mental Disorders; NR, Not reported; NA, Not Applicable; NG, Not given.

**Table 2: Study Characteristics of Non-RCTs**

| **Author (Year)** | **Country** | **No of Participants** | **Age Range** | **Primary clinical characteristics** | **Secondary clinical characteristics** | **Intervention 1/ frequency** | **Intervention 2/ frequency** | **Control** | **Primary Outcome** | **Secondary Outcome** | **Assessment time**  **T1-T7** |
| --- | --- | --- | --- | --- | --- | --- | --- | --- | --- | --- | --- |
| Abdellah et al., 2019 [73] | France | 36 | 18-65 | Adjustment Insomnia and/or history of this disorder within the last 3 months | NR | combination of eschscholtzia and valerian extracts/4 weeks | NA | NA | ISI, Sleep duration, sleep efficiency, sleep latency, | Changes in each ISI components, Anxiety score, VAS, | **Baseline, after 4 weeks** |
| Catherin et al, 2021 [74] | India | 60 | 60–90 years | Stress, anxiety, and sleep quality | NR | Chamomile Oil Aromatherapy/ 3 weeks | NA | NA | PSQI, HAM-A, PSS, | NR | Baseline, 3 weeks |
| Feyzabadi et al., 2014 [75] | Iran | 50 | 16-50 | Chronic insomnia | NR | Viola odorata L. (VO) oil/ 4 weeks | NA | NA | ISI, BDI, Berlin questionnaire | NR | Baseline, Day 15, End of 1 month |
| Cases et al., 2011 [76] | Italy | 20 | 18-70 | mild-to-moderate anxiety disorders | Sleep disturbances | Cyracos® (Melissa officinalis L. leaf extract)/ 2 weeks (15 days) | NA | NA | FRSA, CGI-I, HRSD | NR | Baseline, post-administration, day 15 |
| Kim et al, 2024 [77] | Republic of Korea | 20 | 20-65 years | ISI | Fatigue, daytime sleepiness, and reduced quality of life | Mixture of Rhodiola rosea and Nelumbo nucifera Extracts (RNE)/ 2 Week | NA | NA | ISI, PSQI, Sleep diary parameters | FSS, Short Form-36 Health Survey (SF-36), Safety assessment (vital signs, liver & kidney function tests) | Baseline, 1 week, 2 week |
| Simone et al., 2023 [78] | Italy | 41 | 29-62 | Insomnia Severity | Number of Awakenings Per Night | SPINOFF® fixed combination (Linden, Hawthorn, Vitamin B1, and Melatonin)/ 4 weeks | NA | NA | PSQI, ISI, | Anxiety, stress, | Baseline, 30 days |
| Kimtata et al., 2023 [79] | India | 100 | 18-60 | Mild-moderate stress, anxiety | Insomnia dehydroepiandrosterone (DHEA) | Zero Tension tablet/ 8 weeks | NA | NA | (HAM-A), PSS, ISI, | Hematological parameters, Changes in vital signs, Serum biochemistry | Visit 1 (day 0), Visit 2 (day 2), Visit 3 (day 28, telephonic follow-up), Visit 4 (day 56, end of study) |
| Lemoine et al., 2019 [80] | Poland | 40 | 20-75 years | ISI | NA | Novanuit® Triple Action (combination of melatonin, vitamin B6, and medicinal plant extracts)/ 4 weeks | NA | NA | ISI, Sleep latency, Total sleep time, Sleep efficiency | NA | Baseline, week 3, week 4 |
| Jaroenngarmsamer et al., 2021 [81] | Thailand | 20 | 18-22 | Insomnia / Poor sleep quality | NR | Chamomile essential oil/ 2 weeks | Lavender essential oil/ 2 weeks | NR | Sleep quality score | NR | Baseline, 2 weeks, washover 1 week, 2 weeks |

**Abbreviations:** PSQI, Pittsburg sleep quality Index; PSS, Perceived Stress Scale; ISI, Insomnia severity index; HRSD, Hamilton Rating Scale for Depression; HAM, Hamilton anxiety scale; BAI, Beck Anxiety Inventory; BDI, Beck Depression Inventory; FRSA, Free Rating Scale for Anxiety; SAS, Self-rating Anxiety Scale; ESS, Epworth Sleepiness scale; EPDS, Edinburgh Postnatal depression scale; PFS, Parkinson Fatigue Scale; CGI-I, Clinical Global Impression; M, Mean; SD, Standard deviation; DSM, Diagnostic Statistical Manual of Mental Disorders; VAS, Visual Analogue scale; NR, Not reported; NA, Not Applicable; NG, Not given.

**Table 3: Summary of Continuous Outcomes from RCTs**

| **Study ID** | **Outcome** | **Timing of Outcome** | **Intervention**  **N** | **Intervention Mean** | **Intervention SD** | **Control**  **N** | **Control**  **Mean** | **Control**  **SD** |
| --- | --- | --- | --- | --- | --- | --- | --- | --- |
| Abdullahzadeh et al., 2017 [50] | PSQI | before and 4 weeks after intervention | 40 | 5.05 | ± 3.76 | 37 | 8.24 | ± 4.7 |
| Zick et al., 2011 [51] | PSQI | Week 4 | 17 | 7.5 | ± 3.3 | 17 | 7.1 | ± 2.7 |
| Abdollahzadeh and Naji, 2014 [52] | PSQI | after 8 weeks | 40 | 4.3 | ± 3.7 | 37 | 8.2 | ± 4 |
| Abbasinia et al, 2016 [53] | PSQI | 8 weeks | 55 | 10.1 | ± 1.53 | 55 | 11.02 | ± 2.45 |
| Jarukitsakul et al.,2025 [54] | PSQI | 21 days | 15 | 4.93 | 3.15 | 15 | 5.93 | 3.63 |
| Alvarado-García et al, 2024 [55] | PSQI | 8 weeks | 56 | 3.96 | ±3.35 | 58 | 8.06 | ±5.60 |
| Chang and Chen, 2016 [56] | PSQI | 2 weeks | 35 | 6.11 | 2.867 | 37 | 8.08 | 3.787 |
| Adib-Hajbaghery et al, 2017 [57] | PSQI | 6 weeks | 30 | 9.13 | ± 2.44 | 30 | 11.4 | ± 2.94 |
| Deepa et al., 2024 [58] | PSQI | 2 weeks | 40 | 6.45 | ± 1.65 | 40 | 15.98 | ± 2.15 |
| Renosih et al.,2025 [59] | PSQI | 4 weeks | 30 | 7.8 | 2.5 | 30 | 11.2 | ±2.7 |
| Harit et al., 2024 [60] | PSQI | Week 4 (Day 30) | 32 | 9 | ±5.1188 | 33 | 13.15 | ±4.15 |
| Lee et al,2019 [61] | PSQI | 2 weeks | 45 | 6.91 | ± 1.61 | 39 | 7.05 | ± 1.521 |
| Ngan and Conduit, 2011 [62] | SQ | 1 week | 41 | 3.83 | 0.61 | 41 | 3.57 | 0.56 |
| Marcos et al, 2020 [63] | PSQI | 3 months | 36 | 8.8 | 2.5 | 35 | 9.4 | 2.7 |
| Taavoni et al, 2013 [64] | PSQI | after 4 weeks | 50 | 8.36 | ± 4.46 | 50 | 10.52 | ±2.89 |
| Pierro et al,2024 [65] | ISI | after 14 days of first supplement | 14 | 6.8 | ± 4.1 | 16 | 9.7 | ± 3.7 |
| Ranjbar et al, 2018a [66] | PSQI | 4 weeks | 34 | 7.55 | ± 3.79 | 33 | 10.66 | ±3.55 |
| Ranjbar et al, 2018b [67] | ISI | 4 weeks | 23 | 11.3 | ±5.24 | 22 | 14.31 | ±5.60 |
| Feyzabadi et al, 2017 [68] | PSQI | 4 weeks | 22 | 6.45 | ± 2.30 | 19 | 10.21 | ± 2.93 |
| Taherzadeh et al, 2020 [69] | ISI | 8 weeks | 25 | 13 | ±2.2 | 25 | 14.9 | 2.2 |
| Maroo et al, 2013 [70] | ISI | 2 weeks | 39 | 8.5 | ± 4.64 | 39 | 7.9 | ± 4.03 |
| Gutiérrez-Romero et al, 2024 [71] | PSQI | 6 weeks | 35 | 6.49 | ± 2.5 | 29 | 6 | ±1.7 |
| Bongartz et al., 2019 [72] | mAIS | 6 Weeks | 25 | 11.76 | 6.85 | 25 | 4 | 4.8 |

**Table 4: Results of the continuous outcomes of individual non-RCTs**

| **Study & Intervention** | **Herbal Intervention** | **Outcome Measure** | **Pre-Post Change** | **Interpretation** |
| --- | --- | --- | --- | --- |
| **Single Herbs** | | | | |
| Catherin, 2021 [74] | Chamomile (Matricaria recutita) | Pittsburgh Sleep Quality Index (PSQI) | 10.35-5.55 = 4.8 | LB (Improvement) |
| Feyzabadi et al., 2014 [75] | Sweet Violet (Viola odorata) | Insomnia Severity Index (ISI) | 16.32 -6.48 = 9.84 | B (Large improvement, meets MCID) |
| Cases et al., 2011 [76] | Lemon Balm (Melissa officinalis) | Hamilton Rating Scale for Depression (HRSD) | 1.13 -0.3 = 0.83 | B (Large, stat. significant improvement) |
| Kim et al., 2024 [77] | Lotus & Rhodiola (*Nelumbo nucifera* & *Rhodiola rosea*) | Insomnia Severity Index (ISI) | 12.80 – 10.0 = 2.80 | LB (Modest improvement) |
|  |  | Pittsburgh Sleep Quality Index (PSQI) | 10.25-8.40 = 1.85 | LB (Modest improvement) |
| Jaroenngarmsamer  et al., 2021 [81] | Chamomile (Matricaria recutita) | Sleep Quality Score | 80.20- 55.65 = 24.55 | B (Large, stat. significant improvement) |
| **Multi-Herb Combinations with Confounding Interventions** | | | | |
| Abdellah et al., 2019 [73] | California Poppy (with Valerian) | Insomnia Severity Index (ISI) | 16.09 -11.32 = 4.77 | Not Synthesized (Confounded by Valerian) |
| Simone et al., 2023 [78] | Hawthorn, Linden  (with Melatonin) | Insomnia Severity Index (ISI) | 12.7 - 6.4 = 6.3 | Not Synthesized (Confounded by Melatonin) |
|  |  | Pittsburgh Sleep Quality Index (PSQI) | 8.2 - 6.4 = 1.8 | Not Synthesized (Confounded by Melatonin) |
| Kimtata et al., 2023 [79] | Passiflora (with Valerain) | Insomnia Severity Index (ISI) | 15.1 - 5.5 =9.6 | Not Synthesized (Confounded by Valerian) |
| Lemoine et al., 2019 [80] | California Poppy, Passionflower, Lemon Balm (with Melatonin) | Sleep Quality (0-10) | 5.4 - 7 = -1.6 | Not Synthesized (Confounded by Melatonin) |

Legend: B=Beneficial, LB=Likely Beneficial, NC/U=No Change/Unclear. =Statistically significant (p<0.05) and/or meets the minimal clinically important difference MCID. e.g., a 6–8 point reduction on the ISI).

| **Table 5 :GRADE Summary of findings:** | | | | | | |
| --- | --- | --- | --- | --- | --- | --- |
| **Herbal Sedatives compared to Placebo for Insomnia** | | | | | | |
| **Patient or population:** Insomnia  **Setting:**  **Intervention:** Herbal Sedatives  **Comparison:** Placebo | | | | | | |
| Outcomes | **Anticipated absolute effects^*^** (95% CI) | | Relative effect (95% CI) | № of participants (studies) | Certainty of the evidence (GRADE) | Comments |
|  | **Risk with Placebo** | **Risk with Herbal Sedatives** |  |  |  |  |
| Pittsburgh Sleep Quality Index ( PSQI) Scale from: 3.96 to 10.1 follow-up: range 1 weeks to 8 weeks | - | SMD **0.85 SD lower** (1.38 lower to 0.32 lower) | - | 1134 (16 RCTs) | ⨁⨁⨁◯ Moderate^a,b^ | Herbal Sedatives results in large reduction in Pittsburgh Sleep Quality Index. |
| Insomnia Severity Index (ISI) Scale from: 6.8 to 13 follow-up: range 2 weeks to 8 weeks | - | SMD **0.46 SD lower** (0.93 lower to 0.02 higher) | - | 203 (4 RCTs) | ⨁⨁⨁◯ Moderate^a,b^ | Herbal Sedatives probably reduces Insomnia Severity Index slightly. |
| ***The risk in the intervention group** (and its 95% confidence interval) is based on the assumed risk in the comparison group and the **relative effect** of the intervention (and its 95% CI).  **CI:** confidence interval; **SMD:** standardised mean difference | | | | | | |
| **GRADE Working Group grades of evidence** **High certainty:** we are very confident that the true effect lies close to that of the estimate of the effect. **Moderate certainty:** we are moderately confident in the effect estimate: the true effect is likely to be close to the estimate of the effect, but there is a possibility that it is substantially different. **Low certainty:** our confidence in the effect estimate is limited: the true effect may be substantially different from the estimate of the effect. **Very low certainty:** we have very little confidence in the effect estimate: the true effect is likely to be substantially different from the estimate of effect. | | | | | | |
