## Supplementary material for "Effectiveness of Lesser Known Herbal Sedatives for Insomnia: A Systematic Review and Meta-Analysis": Search string

**Appendix I**

**Search Strategy**

1. **PubMed**

("insomnia"[Title/Abstract] OR "sleep disorder*"[Title/Abstract] OR "sleep disturbance*"[Title/Abstract] OR "Sleep Wake Disorders"[MeSH Terms] OR "sleep initiation and maintenance disorder*"[Title/Abstract] OR "dyssomnia*"[Title/Abstract] OR "poor sleep"[Title/Abstract])

AND

("Passionflower"[Title/Abstract] OR "Passiflora incarnata"[Title/Abstract] OR "Hawthorn"[Title/Abstract] OR "Crataegus oxyacantha"[Title/Abstract] OR "Melissa officinalis"[Title/Abstract] OR "Lemon balm"[Title/Abstract] OR "Chamomile"[Title/Abstract] OR "Matricaria recutita"[Title/Abstract] OR "Sweet Violet"[Title/Abstract] OR "Viola odorata"[Title/Abstract] OR "Lotus"[Title/Abstract] OR "Nelumbo nucifera"[Title/Abstract] OR "Rhodiola"[Title/Abstract] OR "Rhodiola rosea"[Title/Abstract] OR "California Poppy"[Title/Abstract] OR "Eschscholtzia californica"[Title/Abstract])

Results: 169

Dates searched: Jun -July 2025

2. **Euro PMC**

TITLE_ABS:(( "insomnia" OR "sleep disorder*" OR "sleep disturbance*" OR "Sleep Wake Disorders" OR "sleep initiation and maintenance disorder*" OR "dyssomnia*" OR "poor sleep" ) AND ( "Passionflower" OR "Passiflora incarnata" OR "Hawthorn" OR "Crataegus oxyacantha" OR "Melissa officinalis" OR "Lemon balm" OR "Chamomile" OR "Matricaria recutita" OR "Sweet Violet" OR "Viola odorata" OR "Lotus" OR "Nelumbo nucifera" OR "Rhodiola" OR "Rhodiola rosea" OR "California Poppy" OR "Eschscholtzia californica" ))

RESULTS: 181

Dates Searched: Jun -July 2025

3. **Cochrane Library**

(insomnia OR "sleep disorder") AND (passionflower OR "passiflora incarnata" OR hawthorn OR "crataegus oxyacantha" OR "melissa officinalis" OR "lemon balm" OR chamomile OR "matricaria recutita" OR "sweet violet" OR "viola odorata" OR lotus OR "nelumbo nucifera" OR rhodiola OR "rhodiola rosea" OR "california poppy" OR "eschscholtzia californica")

Results: 109

4. **EBSCOhost databases (CINAHL, MEDLINE, APA PsycInfo)**

(TI ( insomnia OR "sleep disorder*" OR "sleep disturbance*" OR "sleep initiation and maintenance disorder*" OR dyssomnia* OR "poor sleep" )

OR AB ( insomnia OR "sleep disorder*" OR "sleep disturbance*" OR "sleep initiation and maintenance disorder*" OR dyssomnia* OR "poor sleep" )

OR SU ( "Sleep Disorders" OR "Sleep Initiation and Maintenance Disorders" OR Insomnia ))

AND

( TI ( passionflower OR "passiflora incarnata" OR hawthorn OR "crataegus oxyacantha" OR "melissa officinalis" OR "lemon balm" OR chamomile OR "matricaria recutita" OR "sweet violet" OR "viola odorata" OR lotus OR "nelumbo nucifera" OR rhodiola OR "rhodiola rosea" OR "california poppy" OR "eschscholtzia californica" )

OR AB ( passionflower OR "passiflora incarnata" OR hawthorn OR "crataegus oxyacantha" OR "melissa officinalis" OR "lemon balm" OR chamomile OR "matricaria recutita" OR "sweet violet" OR "viola odorata" OR lotus OR "nelumbo nucifera" OR rhodiola OR "rhodiola rosea" OR "california poppy" OR "eschscholtzia californica" )

OR SU ( "Plants, Medicinal" OR "Herbal Medicine" OR "Phytotherapy" ))

AND

( TI ( treatment OR therapy OR management OR intervention OR trial OR study OR efficacy OR effectiveness OR safety )

OR AB ( treatment OR therapy OR management OR intervention OR trial OR study OR efficacy OR effectiveness OR safety )

OR SU ( "Treatment Outcomes" OR Management OR Therapy ))

AND

( TI ( "sleep quality" OR "sleep duration" OR "sleep latency" OR "sleep improv*" OR "falling asleep" OR "sleep onset" OR "sleep efficiency" )

OR AB ( "sleep quality" OR "sleep duration" OR "sleep latency" OR "sleep improv*" OR "falling asleep" OR "sleep onset" OR "sleep efficiency" ))

Results: 256

5. **Embase**

('insomnia'/exp OR 'insomnia':ti,ab OR 'sleep disorder*':ti,ab OR 'sleep disturbance*':ti,ab OR 'sleep initiation and maintenance disorder*':ti,ab OR 'dyssomnia*':ti,ab OR 'poor sleep':ti,ab)

AND

('plants, medicinal'/exp OR 'passionflower':ti,ab OR 'passiflora incarnata':ti,ab OR 'hawthorn':ti,ab OR 'crataegus oxyacantha':ti,ab OR 'melissa officinalis':ti,ab OR 'lemon balm':ti,ab OR 'chamomile':ti,ab OR 'matricaria recutita':ti,ab OR 'sweet violet':ti,ab OR 'viola odorata':ti,ab OR 'lotus':ti,ab OR 'nelumbo nucifera':ti,ab OR 'rhodiola':ti,ab OR 'rhodiola rosea':ti,ab OR 'california poppy':ti,ab OR 'eschscholtzia californica':ti,ab)

Results: 136

6. **Google Scholar**

allintitle: (passionflower OR "passiflora incarnata" OR hawthorn OR "crataegus oxyacantha" OR "melissa officinalis" OR "lemon balm" OR chamomile OR "matricaria recutita" OR "sweet violet" OR "viola odorata" OR lotus OR "nelumbo nucifera" OR rhodiola OR "rhodiola rosea" OR "california poppy" OR "eschscholtzia californica") AND (insomnia OR "sleep disorder" OR "sleep disturbance")

Results: 50

7. **Directory of Open Access Journals (DOAJ)**

abstract:(passionflower OR "passiflora incarnata" OR hawthorn OR "crataegus oxyacantha" OR "melissa officinalis" OR "lemon balm" OR chamomile OR "matricaria recutita" OR "sweet violet" OR "viola odorata" OR lotus OR "nelumbo nucifera" OR rhodiola OR "rhodiola rosea" OR "california poppy" OR "eschscholtzia californica") AND (insomnia OR "sleep disorder" OR "sleep disturbance")

Results: 14

8. **ClinicalTrials.gov**

Condition: insomnia OR "sleep disorder" OR "sleep disturbance" OR "sleep initiation and maintenance disorder" OR dyssomnia OR "poor sleep"

Intervention: passionflower OR "passiflora incarnata" OR hawthorn OR "crataegus oxyacantha" OR "melissa officinalis" OR "lemon balm" OR chamomile OR "matricaria recutita" OR "sweet violet" OR "viola odorata" OR lotus OR "nelumbo nucifera" OR rhodiola OR "rhodiola rosea" OR "california poppy" OR "eschscholtzia californica"

Result: 10

9. **International Clinical Trials Registry Platform (ICTRP)**

insomnia OR "sleep disorder" OR "sleep disturbance" OR "sleep initiation and maintenance disorder" OR dyssomnia OR "poor sleep"

AND

passionflower OR "passiflora incarnata" OR hawthorn OR "crataegus oxyacantha" OR "melissa officinalis" OR "lemon balm" OR chamomile OR "matricaria recutita" OR "sweet violet" OR "viola odorata" OR lotus OR "nelumbo nucifera" OR rhodiola OR "rhodiola rosea" OR "california poppy" OR "eschscholtzia californica"

Results: 2
